# Distinct lung cell signatures define the temporal evolution of diffuse alveolar damage in fatal COVID-19

**DOI:** 10.1101/2023.05.05.23289594

**Authors:** Luke Milross, Bethany Hunter, David McDonald, George Merces, Amanda Thompson, Catharien M.U. Hilkens, John Wills, Paul Rees, Kasim Jiwa, Nigel Cooper, Joaquim Majo, Helen Ashwin, Christopher J.A. Duncan, Paul M. Kaye, Omer Ali Bayraktar, Andrew Filby, Andrew J. Fisher

## Abstract

**Background:** Lung damage in severe COVID-19 is highly heterogeneous however studies with dedicated spatial distinction of discrete temporal phases of diffuse alveolar damage (DAD) and alternate lung injury patterns are lacking. Existing studies have also not accounted for progressive airspace obliteration in cellularity estimates. We used an imaging mass cytometry (IMC) analysis with a novel airspace correction step to more accurately identify the cellular immune response that underpins the heterogeneity of severe COVID-19 lung disease.

**Methods:** Lung tissue was obtained at post-mortem from severe COVID-19 deaths. Pathologist-selected regions of interest (ROIs) were chosen by light microscopy representing the patho-evolutionary spectrum of DAD and alternate disease phenotypes were selected for comparison. Architecturally normal SARS-CoV-2-positive lung tissue and tissue from SARS-CoV-2-negative donors served as controls. ROIs were stained for 40 cellular protein markers and ablated using IMC before segmented cells were classified. Cell populations corrected by ROI airspace and their spatial relationships were compared across lung injury patterns.

**Results:** Forty patients (32M:8F, age:22-98), 345 ROIs and >900k single cells were analysed. DAD progression was marked by airspace obliteration and significant increases in mononuclear phagocytes (MnPs), T and B lymphocytes and significant decreases in alveolar epithelial and endothelial cells. Neutrophil populations proved stable overall although several interferon-responding subsets demonstrated expansion. Spatial analysis revealed immune cell interactions occur prior to microscopically appreciable tissue injury.

**Conclusions:** The immunopathogenesis of severe DAD in COVID-19 lung disease is characterised by sustained increases in MnPs and lymphocytes with key interactions occurring even prior to lung injury is established.

## 1. Introduction

Refractory respiratory failure is the leading cause of death in critically ill COVID-19 patients^1^ and immune-mediated acute lung injury rather than direct cytotoxic effects of SARS-CoV-2 infection appears central to disease severity^2^. Characterisation of the host immune response at the lung tissue level using multimodal approaches is critical to understanding disease pathophysiology^3^.

Diffuse alveolar damage (DAD) is the predominant histological pattern in post-mortem lung tissue (PMLT) from cases of acute severe COVID-19^4^ and is considered a characteristic feature. DAD is routinely divided by pathologists into an acute exudative phase (EDAD), which may progress to a proliferative and organising phase (ODAD)^5^. DAD stages frequently coexist within a single patient^6^ as a temporally heterogeneous pathology. Alternate patterns of lung tissue damage are also recognised^7^, including superimposed bacterial bronchopneumonia (BRON)^5^, pulmonary oedema consistent with acute cardiac failure (PO-ACF)^8^ and invasive pulmonary mycoses (IPM)^9^. How these different phases of DAD and the alternate patterns of lung injury influence the nature of the immune response is currently unclear.

Immune-mediated acute lung injury rather than the direct cytotoxic effects of SARS-CoV-2 infection itself appears to be central to severe or fatal COVID-19, evidenced by a topological dissociation between inflammatory and viral-positive areas^2^, reduced or absent virus in late disease^10,11^ and that therapeutically, the inflammation-modulating glucocorticoid dexamethasone provides a significant mortality reduction in severe disease^12^. The systemic immune response in COVID-19 shows major shifts in lymphoid and myeloid compartments as blood signatures of severe disease^13^ and the ‘competent’ immune profiles associated with mild COVID-19^14^. However, these studies provide mere inferences to the cellular responses and architectural injury hidden at the tissue level where the end organ dysfunction occurs and detailed immunophenotyping of affected tissue is required to complete the picture^15–17^.

A suite of advanced pathology techniques were utilised early in the pandemic to dissect the shifts in immune and structural cells in COVID-19 post-mortem lung tissue (PMLT)^3^. Major emerging themes included a significant macrophage infiltration, expansion of T and B lymphocytes and mesenchymal and fibroblastic proliferation^15,18^ and intriguingly a topological dissociation between inflammatory and viral-positive areas^2^. Published analyses of COVID-19 PMLT have significant limitations due to indiscriminate comparison of control tissue with an undifferentiated amalgam of COVID-19 ‘diseased tissue’^3^. Furthermore, the metric commonly used to quantify immune cells in lung tissue, namely *cells per unit area of tissue section*, generally expressed as ‘cells/mm^2^’^15,18–20^ can be confounded by changes in airspace contributions to section area.

In this paper, we studied lung tissue from a cohort of 40 people who died with severe COVID-19 and analysed pathologist-guided lung tissue regions of interest (ROIs) representing the distinct temporal stages of DAD as well as alternate COVID-19-related disease patterns.

## 2. Methods

### Tissue Bank Assembly and Cohort Description

Lung tissue was obtained via autopsy with next of kin consent from three United Kingdom Biobanks (see Supp Methods) from individuals with clinical and microbiological evidence that COVID-19 disease was the primary precipitant of death. All patients were confirmed SARS-CoV-2-positive by reverse transcription polymerase chain reaction (RT-PCR) of nasopharyngeal and/or direct lung tissue swabs at autopsy. We included a cohort of 40 adults, whose clinical metadata was obtained from electronic medical records and post-mortem reports (Figure 1, Supp Tables 1-3). Architecturally preserved control tissue, henceforth referred to as ‘PRESneg’ was obtained from unused lung donors which did not proceed to transplantation (n=2).

**Figure 1:**
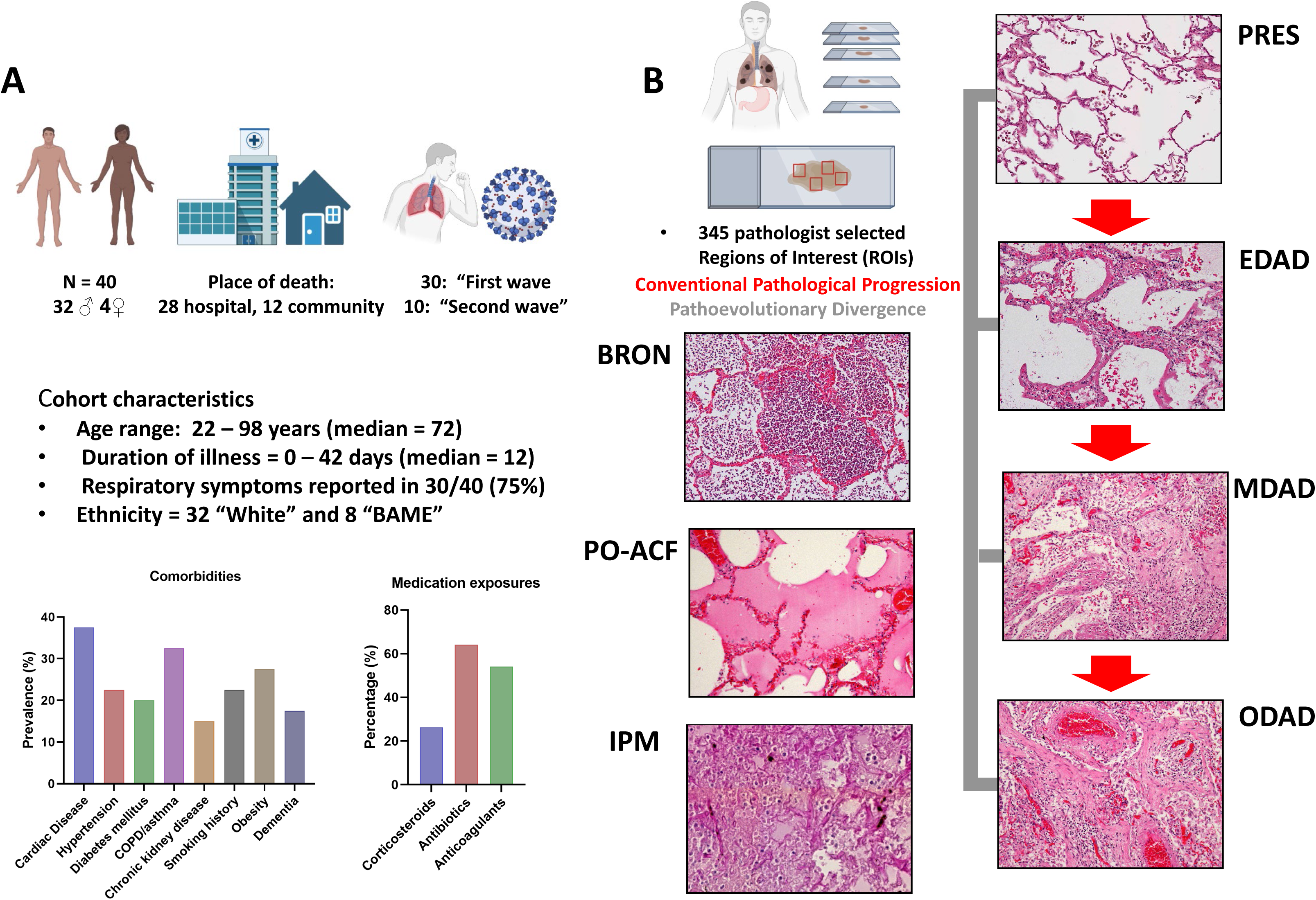
Overview of the cohort demographics and histological model. A) A graphical summary of the cohort composition and key clinical metadata, including comorbidities and exposure to medication. B) A graphical and histological summary of the different pathology states present in the cohort as identified by expert pathologist input (H&Es at X100 magnification except for IPM which is at X200 magnification). Note that the conventional progression stages are shown with red directional arrows and that the pathoevolutionary divergent stages are linked by grey lines to denote that progression and origin are unknown with respect to conventional stages of pathology. Histology images are derived from H&E stained FFPE serial tissue sections adjacent to those used for IF and IMC analysis. Please also note that “PRES” pathology falls in to two classes; derived from SARS-CoV2 infected and uninfected tissue.

### Tissue Section Preparation and ROI Selection

Formalin-fixed paraffin-embedded (FFPE) lung tissue blocks from multiple lung regions were serially cut and mounted onto slides (Supp Methods). The H&E-stained primary slide from each serial deck was scanned onto the open-source digital online pathology platform OMERO (‘The Open Microscopy Environment’). ROIs were selected under guidance of a consultant histopathologist with cardiothoracic expertise with sizes ranging from 0.25mm^2^ (500µm x 500µm) to 1mm^2^ (1000µm x 1000µm). ROI classifications included the temporal phases of DAD, bronchopneumonia (‘BRON’), pulmonary oedema consistent with acute cardiac failure (‘PO-ACF’) and invasive pulmonary mycosis (IPM). ROIs were also selected from DAD-free, ‘preserved’ regions of lung tissue from these SARS-CoV-2 infected individuals (titled PRESpos), as well as from SARS-CoV-2 negative patients (PRESneg).

DAD was divided based on published histological criteria^21^ into exudative DAD (EDAD), organising DAD (ODAD) and mixed (or ‘intermediate’) diffuse alveolar damage (‘MDAD’). Of these criteria, selection was weighted by a primary ‘hallmark’ characteristic in the context of secondary supportive features (Supp Table 4).

### Manufacture of Control Tissue MicroArray (TMA) Material

To provide positive and negative staining controls for all 40 antibodies in our panel as well as provide empirical controls for batch effects, we prepared FFPE Tissue MicroArray (TMA) blocks composed of human tonsil tissue and SARS-CoV-2-infected and mock infected Vero E6 cells. Detailed exploration of TMA manufacture is provided in Supp Methods.

### Antibody panel design, conjugation and antigen retrieval for Imaging Mass Cytometry Analysis

The 40-plex antibody panel identifying the immune, signalling and stromal components in the surrounding microenvironment of COVID19 post-mortem lung tissue is described in Supp Table 5. All antibodies used in this study were pre-validated for performance using Tris EDTA pH9 “Heat-Induced Epitope Retrieval” (HIER) two colour immuno-fluorescence (IF) and conjugated (where necessary) to lanthanide metals and fully validated as described in Supp Methods.

### Hyperion (IMC) set up, quality control (QC) and sample acquisition

The PM tissue cohort was acquired using a fully calibrated and quality controlled Hyperion Tissue Imager over 12 individual batches with a TMA control slide processed and stained alongside (see Supp Methods). Ablations were performed at 200Hz laser frequency creating MCD files containing all data from a given ROI for each slide/case. MCD files were analysed in MCD Viewer software (Standard Bio-Tools) to perform a qualitative QC of the staining intensity and pattern against the benchmark of IF validation images. All images were exported as 16-bit single multi-level TIFFs. The multi-level 16-bit TIFF images were then input into the OPTIMAL pipeline^22^ for data exploration at the single cell, spatial level.

### Cell segmentation, feature extraction, parameter correction/normalisation and FCS file creation

Cell segmentation was performed as per the OPTIMAL pipeline^22^ using Ilastik. Output nuclear probability maps were input into CellProfiler to segment cell nuclei, that in turn acted as seeds for cell segmentation using a propagation algorithm based on the EPCAM signal (Supp Figure 1). Single cell objects were measured for mean intensity in each of the labelled channels and corrected for metal signal “spillover” according to a previously described approach^23^. An *arcsinh* transformation cofactor (c.f.) of 1 was applied to all metal signal parameters. Batch effect correction was performed using Z-score normalisation on the *arcsinh* c.f. 1 transformed data (Supp Figure 2). We also added additional metadata to the files such as batch number, to be accessible and plot-able parameters for subsequent analysis. Final matrix data was converted to .FCS file format within the MATLAB pipeline.

### Visualisation, clustering and spatial exploration of single cell IMC data

FCSExpress software (Version 7.14.0020 or later, Denovo software by Dotmatics, USA) was used as outlined in the OPTIMAL method^22^. Briefly, the FCS files created from the segmentation pipeline of each ROI were loaded as a single merged file. Gates were created on batch, biobank source, pathology class etc. to aid with meta-analysis. SARS-CoV-2 spike and nucleocapsid protein expression was determined for each of the 8 pathology classes on a per cell basis using histogram displays, and the population means were compared to the SARS-CoV-2 infected and mock infected Vero E6 cell TMA controls. Single cell data structure for all 38 positive signals (*arcsinh* c.f. 1 transformed and Z score normalised) was displayed by creating a PacMap dimensionality reduction plot as described previously^22^. To identify resident cell types and states the same 38 transformed and normalised metal parameters were used as input in to the FLOWSOM clustering algorithm as outlined previously^22,24^. The default 100 SOMs (clusters) were merged using a hierarchical approach to 40 consensus SOMs (cSOMS). The 40 cSOMs were further merged to 25 final “tier 2” clusters based on expert annotation and *a priori* knowledge from heat map interrogation. “Tier 2” clusters were then merged to 10 “high level” cell types denoted as “tier 1”. Spatial neighbourhood analysis for tier 1 and tier 2 clusters was performed as outlined in Hunter et al. (disc outgrowth of 5 pixels and 100 iterations of randomly mapping cells back on to the segmentation maps)^22^. Statistically significant interactions between cell types were determined by comparing spatial cell iterations and those obtained by the random permutations of the cell positions. If differences were detected in the original data compared to a 90% confidence interval of the random iterations, then a significant difference (interaction or avoidance) was listed for that cell type. These positive, neutral, and negative interactions were then averaged to create the overall heatmap for the condition (i.e., pathology, region, etc.). These interactions were assessed across all 8 pathology classes.

### Airspace correction, normalisation of cell counts by tissue area and statistical analysis

The metric commonly used to quantify immune cells in lung tissue, namely *cells per unit area of tissue section*, generally expressed as ‘cells/mm^2^’^15,18–20^ can be confounded by changes in airspace contributions to section area. To compensate for the confounding effect of airspace obliteration, we developed a correction method to standardise cellularity when describing distinct lung cell signatures in COVID-19 PMLT. For each ROI, the percentage of airspace was determined using the following equation (1):

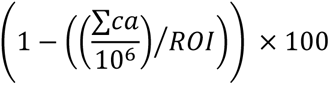

Where *ca* = the area of each single cell in each ROI in µm^2^ and *ROI* = the area of the imaged ROI in mm^2^. Furthermore, the area of cellular tissue in mm^2^ (first part of equation 1 above) was used to normalise all cell counts to account for any artificial increases due to tissue collapsing into the imaged ROI due to loss of air gaps (henceforth referred to as *airspace correction*). For differential analysis of cell counts and percentages at tier 1 and tier 2, the Kruskal–Wallis one way analysis of variance test was performed in GraphPad Prism version 9.5.0 with results considered statistically significant at p<0.05.

## 3. Results

### Demographics and clinical features

Lung tissue was analysed from 40 individuals who died with severe COVID-19 (8F/ 32M). Age at death was 22 – 98 years (median=72 years), with 28/40 (70%) dying in hospital and 12/40 (30%) dying in the community. Duration of illness ranged from 0-42 days (median=12 days, duration of illness data known in 35/40 cases). 30/40 (75%) died in the ‘first wave’ of the pandemic (before 1^st^ October 2020) and 10/40 (25%) died in the ‘second wave’ defined by increasing predominance of the alpha (B.1.1.7) variant^25^. None were vaccinated against SARS-CoV-2. Autopsy was performed same-day to a maximum of seven days (median=3 days) after death. All were SARS-CoV-2 positive by RT-PCR on pharyngeal or direct lung sampling. Supp Tables 1 and 2 and Figure 1 show cohort demographics, comorbidities and disease characteristics. Spike and nucleocapsid proteins have previously been detectable in COVID-19 PMLT analysed by IMC^15^. However, using single cell expression data in positive and negative infected cultured cells as controls, we did not detect spike or nucleocapsid protein in any of the pathology phenotypes (Supp Figure 3).

### Histopathological assessment reveals pathological heterogeneity

Regions of interest (ROIs) were selected representing the temporal phases of DAD and alternate pathological patterns as shown in Figure 1. DAD was the commonest histopathological phenotype, identified in 29/40 cases (72.5%). Of these, 17 patients (58.6%) showed DAD in different evolutionary phases, indicating significant intra-patient temporal heterogeneity. 7 cases were predominantly BRON, 2 cases PO-ACF and one case IPM. A total of 345 ROIs were selected for analysis. The number of ROIs selected in each pathological pattern is found in Supp Table 3.

### Single cell analysis reveals that airspace obliteration not increased cellularity, defines DAD progression

345 ROIs, covering ~195mm^2^ tissue area, were ablated by imaging mass cytometry and single cell analysis was performed using the OPTIMAL^22^ approach (Figure 2A, Supp Figure 1). The pipeline included normalisation for batch effect from both run and tissue source (Supp Figure 2). This process generated a total output of ~901k single cells (Figure 2B).

**Figure 2:**
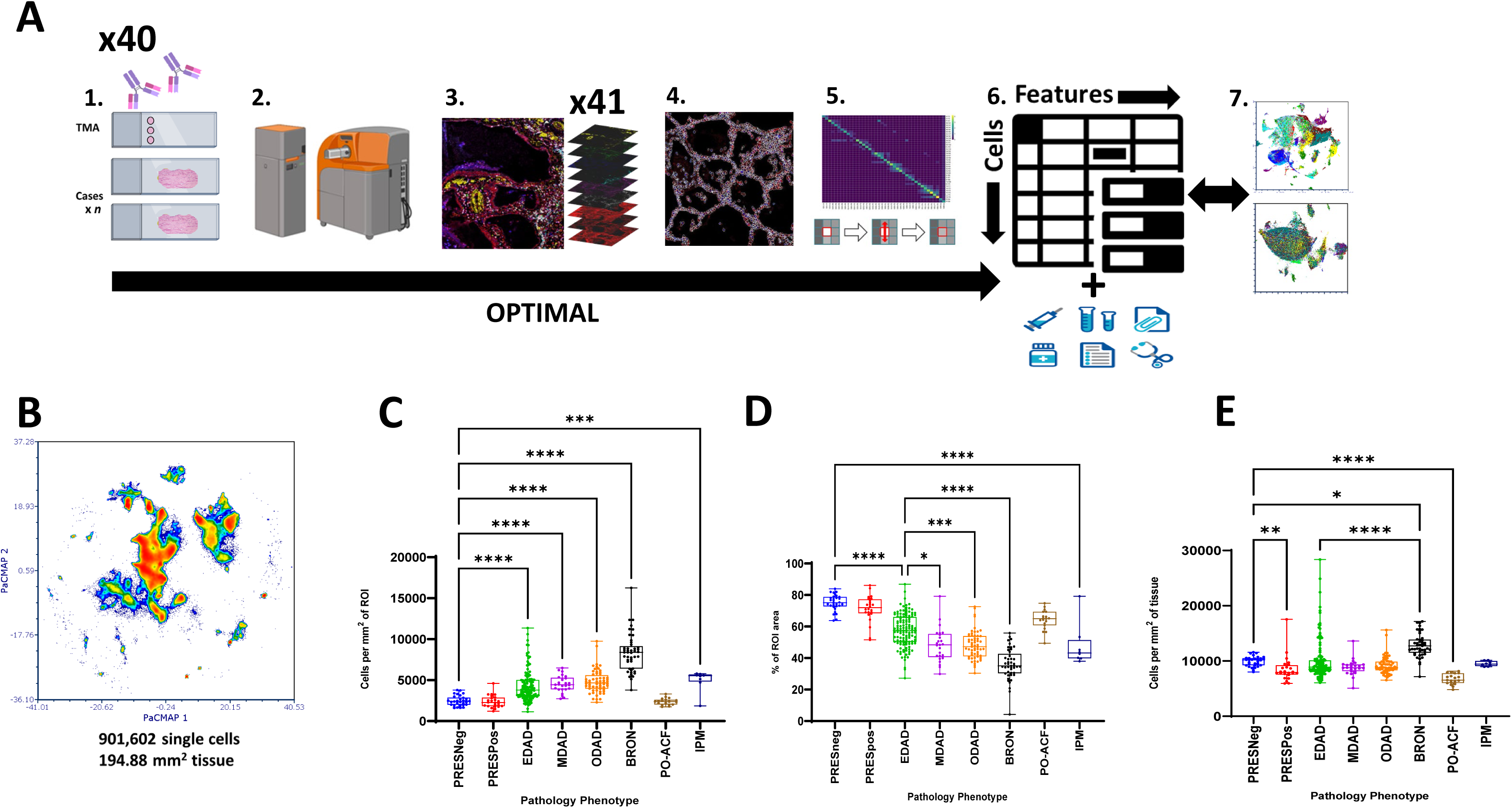
OPTIMAL analysis of single cells in COVID19 PM lung tissue reveals a progressive loss of lung air space leading to elevated cellularity due to tissue obliteration rather than a *de* novo cellular influx. A) PM lung tissue slides were stained with a panel of 40 metal tagged antibodies alongside an additional control TMA slide (1). The pathologist-marked ROIs were then set using an OMERO reference image and ablated using a Hyperion IMC system (2) to produce a set of 41 multispectral images (3) that were segmented to single cell data (4), corrected for spill-over and other factors that could affect clarity (5) and converted to .FCS file format with additional key metadata added (6). Batch effect was determined and corrected for using a z-score normalisation approach (7). B) A PacMap dimensionality reduction plot for all 901,602 single cells representing ~195 mm^2^ of COVID19 PM lung tissue. C) Cell counts per mm^2^ of the ablated ROI area for each of the 8 pathology classes. D) A graph showing the % of air space within each ROI as a function of pathology class. E) A graph of the cell counts per mm^2^ of actual lung tissue in each ROI. Differences between pathology classes were considered statistically significant at p<0.05.

Cellularity was then assessed across each pathology type using total cells normalised to area of each ROI (Figure 2C). There were significant increases in cellularity as the temporal phases of DAD progressed from preserved tissue, and BRON had the highest. However, it was unclear whether increased cellularity in DAD was related to actual cellular influx, or airspace obliteration leading to an increase of tissue within the ROI, or a combination of both. For example, Figure 2D demonstrates striking loss of airspace across DAD progression. To account for this, we normalised cell counts by actual cellular tissue area by airspace correction, as opposed to ROI area. Figure 2E demonstrates the effect of airspace correction in nullifying the increased cellularity previously seen across DAD, indicating that this was confounded by airspace obliteration.

### Increases in mononuclear phagocytes and lymphocytes and not neutrophils define the immune signature of COVID-19 DAD progression

Our high-level (Tier 1) analysis of immune and structural cells generated 10 consensus clusters (Figure 3A, Supp Figure 4), which were substantially discrete when mapped back to a PacMap dimensionality reduction plot (Figure 3B).

**Figure 3:**
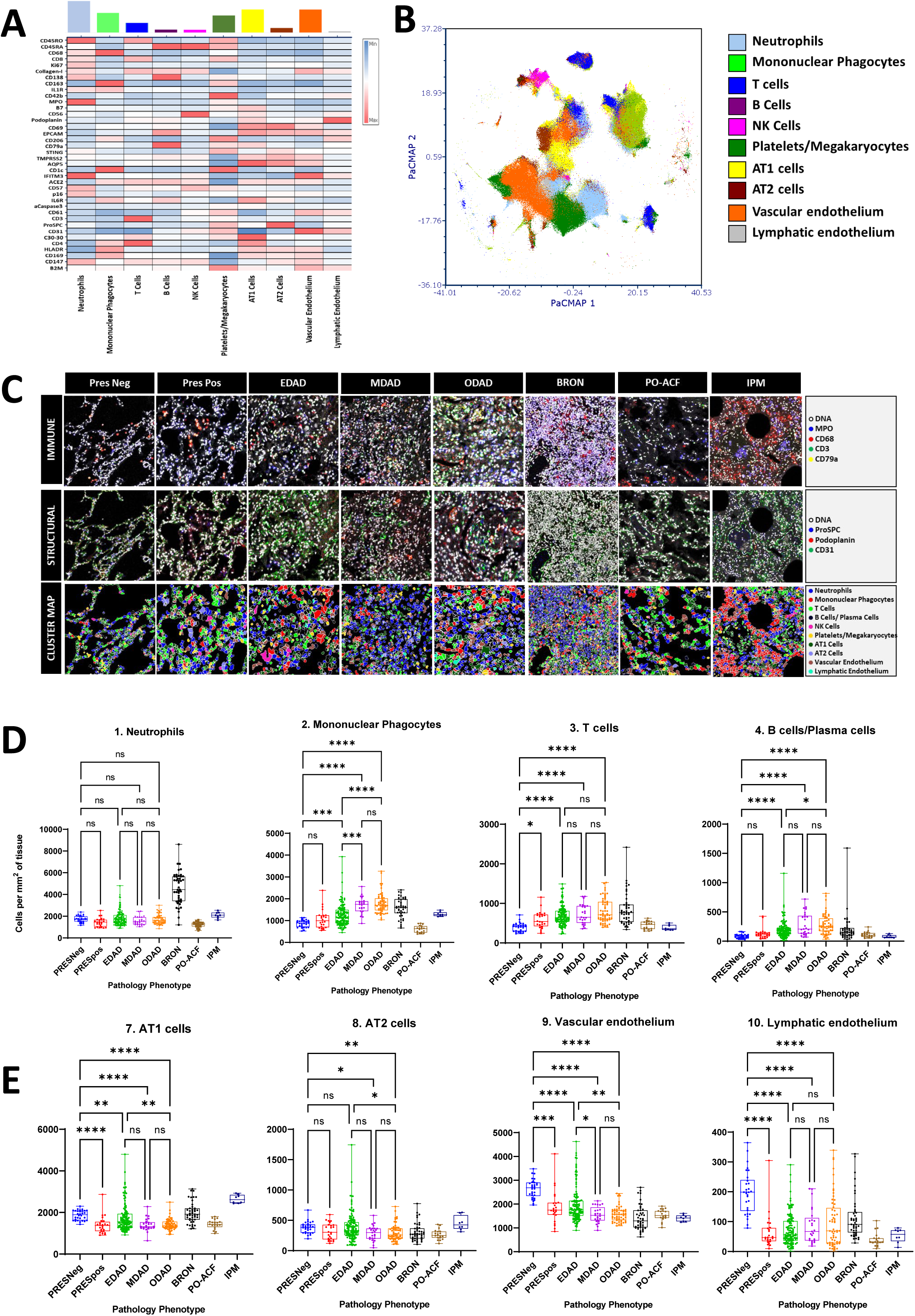
Analysis of Tier 1 cell type clusters reveals key immune and structural cell signatures defining temporal stages of DAD and alternate pathology classes. A) A heat map showing the median Z-score normalised values for all 38 phenotypic and functional markers (SARS-CoV2 Spike and Capsid were deemed to be negative so not used) for tier 1 level clusters. The coloured bars denote the frequency of each cluster across the entire single cell data set. B) A PacMap of the single cell level data coloured by tier 1 cluster as shown in the associated legend. C) Upper panels show pseudo-coloured overlaid key immune cell markers for representative ROIs from all 8 pathology classes. Middle panels show pseudo-coloured overlaid key structural cell markers for the same 8 representative ROIs. Lower panels show cluster maps for the same 8 representative ROIs with all 10 tier 1 clusters. D) Graphs showing the cell counts per mm^2^ of lung tissue per ROI for key tier 1 immune cell types. From left to right; neutrophils, mononuclear phagocytes, T cells and B/Plasma Cells. E) Analogous graphs as shown in D but for major tier 1 structural cell types. From left to right; AT1, AT2, vascular endothelium and lymphatic endothelium. Differences between pathology classes were considered statistically significant at p<0.05.

We analysed pathology phenotypes for their Tier 1 immune cell airspace-corrected cellularity (Figure 3D-E) and proportions (Supp Figure 5A). Neutrophils, alongside a modest rise in mononuclear phagocytes, were seen in BRON but not DAD.

Mononuclear phagocyte and lymphocyte infiltration represent the predominant immune cell hallmarks of COVID-19 DAD, with both showing significantly increased proportions and airspace-adjusted cellularity as DAD evolved (Figure 3C-D). Lymphocyte increases involved both CD4+ and CD8+ T cells and B-cells/plasma cells. Analysis of neutrophils with cells/mm^2^ tissue *prior to* airspace correction was misleading, as this metric suggested a significant increase in DAD classes compared to PRESneg (Supp Figure 6). However, when applying proportion and airspace-adjusted cellularity metrics, no significant differences in neutrophils were seen across any of the temporal phases of DAD and when compared with preserved tissue.

Subgroup analysis was next performed with a focus on EDAD ROIs, with data points coded by sex, ethnicity and pandemic wave. When our PacMap dimensionality reduction plot was coloured for these demographic differences, no obvious difference in data spread was seen (Supp Figure 7). Comparisons between airspace-corrected cellularity were made for neutrophils, MnPs, T cells and B cells/plasma cells coded for sex, ethnicity and pandemic wave demonstrated only subtle differences including significantly increased neutrophils and T cells in second wave compared to first wave EDAD ROIs and significantly increased T cell infiltration in male compared to female EDAD ROIs.

### Progressive loss of alveolar epithelial cells (AECs), vascular endothelial and lymphatic endothelial cells is seen in progressive COVID-19 DAD

Significant decreases in AT1 cells from PRESneg to EDAD and from EDAD to ODAD were identified (Figure 3E). AT2 cells were generally stable in proportion with respect to preserved tissue however there was a significant decrease in MDAD and ODAD compared to PRESneg. Vascular endothelial cells and lymphatic endothelium decreased as DAD progressed. Figure 3C shows raw IMC images and cluster maps for Tier 1 populations in all pathology classes, visually displaying variations in immune and structural populations.

### Individual immune cell phenotypes characterise temporal phases of the COVID-19 DAD continuum

We next used a 38-marker panel with the FLOWSOM algorithm to identify 25 “Tier 2” clusters) and used a Z score normalised heat map to annotate the cell types and states (see Figure 4A). We then focused on the PRES and DAD groups and the airspace-corrected cellularity metric. Supp Figures 8-10 contain analyses by other metrics.

**Figure 4:**
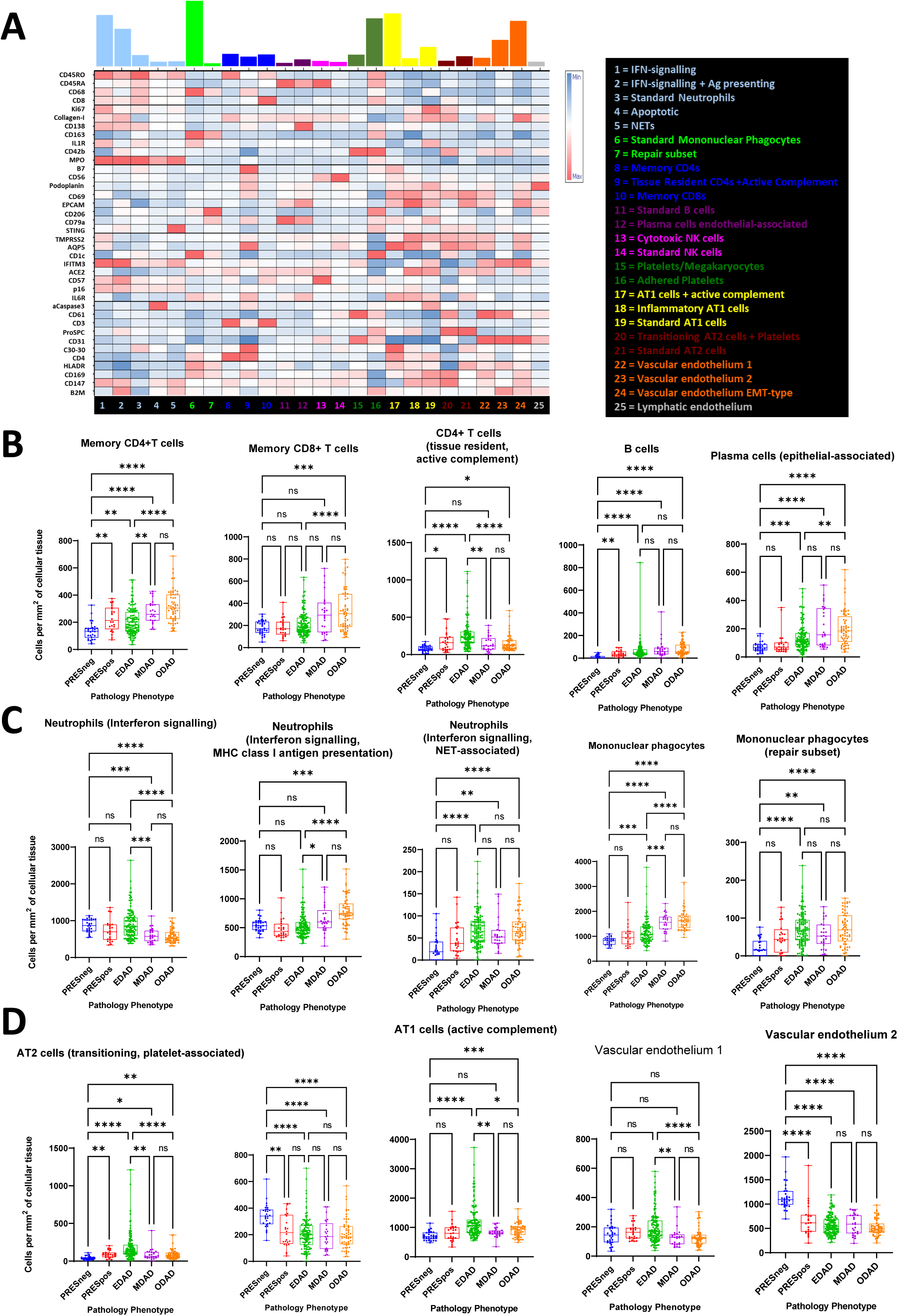
Tier 2 cluster analysis reveals a unique set of cell signatures linked to DAD progression. A) A heat map showing the median Z-score normalised values for all 38 phenotypic and functional markers (SARS-CoV2 Spike and Capsid were deemed to be negative so not used) for tier 2 level clusters. The coloured bars denote the frequency of each cluster across the entire single cell data set. The cluster ID is given by the number below the column (cluster) and denoted in the legend. B) Graphs showing the cell counts per mm^2^ of lung tissue per ROI for key tier 2 lymphocytic cell types. From left to right; memory CD4 T cells, memory CD8 T cells, CD4 T cells with active complement and tissue resident features, B cells and epithelial-associated plasma cells. C) Graphs showing the cell counts per mm^2^ of lung tissue per ROI for key tier 2 non-lymphocytic immune cell clusters. From left to right; neutrophils with signatures of interferon signalling and MHC-class 1 presentation, neutrophils with interferon signalling and NET-associated signatures, mononuclear phagocytes and mononuclear phagocytes with a repair-promoting signature. D) As in B and C but showing key tier 2 structural cell clusters. From left to right; AT2 cells with a transitioning and platelet associated signature, AT 2 cells, AT1 cells with active complement, vascular endothelium 1 (CD61+ CD31+ cells mapping to capillaries) and vascular endothelium 2 (CD61+ CD31+ cells mapping to vascular structure outlines of greater size than capillaries). Differences between pathology classes were considered statistically significant at p<0.05.

With respect to adaptive immune cells (Figure 4B), the significant rises in lymphocytes were accounted for by increased memory CD4+ T cells, memory CD8+ T cells, CD4+ T cells, B cells and plasma cells. Plasma cell infiltration was particularly marked as DAD progressed. Although total neutrophil infiltration is not a hallmark of COVID-19, two phenotypic subsets of neutrophils characterised by interferon signalling (IFITM3^HI^ and STING^HI^), MHC class I antigen presentation (beta-2 microglobulin^HI^) and neutrophil-extracellular traps were significantly increased in as DAD progressed as shown in Figure 4C. An inflammatory subset of mononuclear phagocytes (IL1R^HI^, IL6R^HI^ and HLA-DR^HI^) also significantly increased from PRESneg to EDAD and again from EDAD to ODAD. A second cluster of macrophages, phenotypically consistent with M2 polarisation (CD206^HI^) were increased from PRESneg to DAD phases. No further increases were noted between EDAD, MDAD and ODAD, perhaps suggesting an exhausted reparative process.

Amongst the structural clusters (Figure 4D), we observed an AT1 and a CD4+ T cell cluster with markers suggestive of complement activity (C30-30^HI^ and B7^HI^) significantly elevated in EDAD compared to PRESneg and ODAD. SARS-CoV-2 can activate the complement system via the classical, lectin and alternative pathways or indirectly through endothelial injury and thromboinflammation^26^ and our results suggests an association with AT1 cells and a CD4+ T cells especially during early DAD. A subset of AT2 cells is seen falling as disease progresses which may indicate a known transition process to an AT1 phenotype to replace those lost in the tissue^27^.

### Critical immune cell interactions are established early, prior to overt tissue damage

Analysis of cellular neighbourhoods at Tier 1 immune cell level is showed in interaction/avoidance heat maps for PRES and DADs (Figure 5). All other pathologies are shown in Supp Figure 11 for Tier 1 and in Supp Figure 12 for Tier 2. Marked differences occurred between PRESneg and PRESpos, suggesting critical interactions are established early, prior to overt tissue damage. Notably, in PRESpos, neutrophils appear to interact more with AT2 cells, the primarily infected cell in lung tissue in previous literature^28^, and less with AT1 cells. The most striking specific difference was an increased interaction between neutrophils with interferon signalling and MHC class I antigen presentation markers and AT2 cells (Supp Figure 12). Additionally, we noted mononuclear phagocytes interacted more with both neutrophils and T cells consistent with innate-adaptive crosstalk. Other marked interactions included neutrophils (interferon signalling) and mononuclear phagocytes with inflammatory markers (IL1R^HI^, IL6R^HI^ and HLA-DR^HI^) (Supp Figure 12). Compared to PRESneg tissue, PRESpos tissue showed increased interactions between CD4+ T cells or memory CD8+ T cells and a repair subset of macrophages (CD206^HI^) (Supp Figure 12). M2 macrophages are known for their roles in tissue repair and reduce inflammation via suppression of effector T cells^29^, indicating this process may start early, prior to overt tissue damage. Finally, B cells seemed to interact more with vascular endothelial cells in PRESpos compared to PRESneg, which may indicate early diapedesis and recruitment of antibody producing cells.

**Figure 5:**
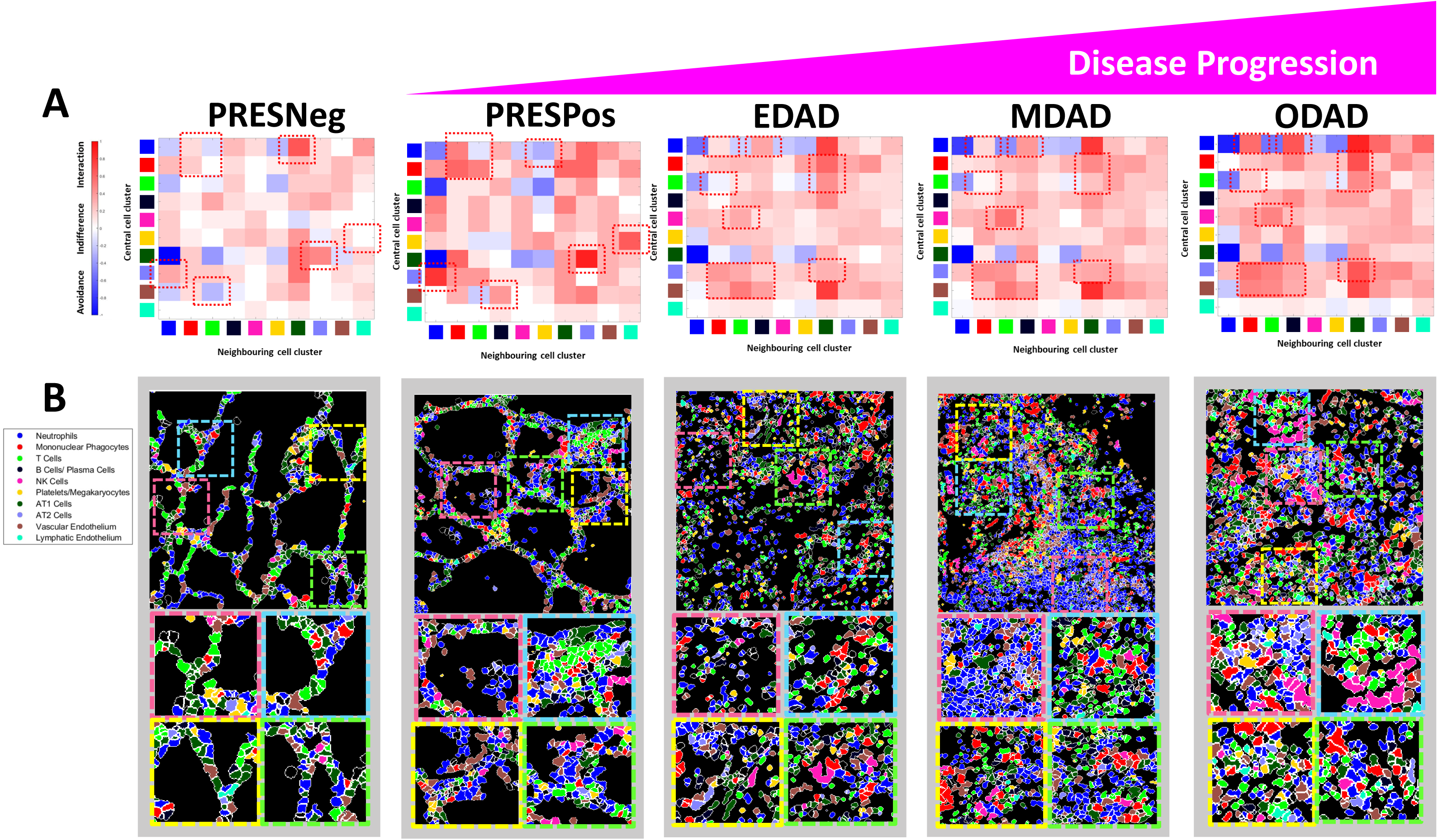
Spatial neighbourhood analysis of tier 1 clusters reveals key interactions and avoidances that correlate with DAD initiation and progression. A) Heat maps for each of the 5 major pathology classes involved in classical disease progression showing the significance of interaction, avoidance or indifference for all 10 tier 1 clusters as per the legend. Red dotted squares are shown to aid with identifying and interpreting key interactions or avoidances. B) Representative cluster maps with coloured boxes denoting areas of focus below whereby example of interactions from A are shown.

A more consistent interactome existed when comparing the DAD tissue phenotypes, however several notable changes were observed including 1) B cells/plasma cells increasing their interactions with neutrophils and mononuclear phagocytes, 2) T cells increasing their interactions with NK cells and 3) mononuclear phagocytes increasing their interactions with multiple cell types including neutrophils and AT2 cells and vascular endothelium.

## 4. Discussion

In this study, we present a comprehensive assessment of the immune cell signature and structural cell composition in lung tissue from fatal COVID-19. Our data places particular emphasis on spatial and temporal differences in the heterogeneous patterns of tissue injury. We show that the pathological evolution of DAD, the archetypal lung pattern in COVID-19, is characterised by sustained increases in mononuclear phagocytes and lymphocytes without a shift in overall neutrophil counts but with shifts in functional neutrophil subsets. Within the structural compartments, there is a loss of AECs and endothelial cells. We confirm that significant airspace obliteration accompanies DAD progression and show this is a significant confounder in measuring relative and absolute cellularity in diseased lung tissue. Finally, critical immune and structural cell interactions occur prior to overt tissue injury.

The major immune cell shifts detected in the COVID-19 lung have been significant macrophage infiltration, expansion of T and B lymphocytes and mesenchymal and fibroblastic proliferation^15,18^. However, interpretation of high resolution molecular pathology studies on COVID-19 PMLT has, to date, been limited by lack of discrimination between histologically different regions of interest^3^ in highly heterogeneous tissue^7–9,30^. Rather, COVID-19 PMLT has hitherto been classified as ‘early’ or ‘late’ disease by chronological duration of illness^15^, by comparison of viral negative and viral positive areas^31,32^, by provision of an inflammation severity score^2^ or simply being compared generally/collectively with non-infected control tissue^19^. Our approach is instead based on the temporal phases of COVID-19 DAD evolution rooted in standardised pathological terminology first established by Katzenstein *et al.* in the 1970’s^33^. Erjefalt *et al.* (2022), using a multiplex immunohistochemistry platform also used the approach of discriminating between exudative, intermediate and organising DAD^18^. Their results are consistent with our findings showing macrophages, B cells and both CD4+ and CD8+ T lymphocytes gradually increasing as DAD progresses.

Our findings confirm that mononuclear phagocyte infiltration is a major hallmark of COVID-19 lung tissue, proven across multiple modalities^2,15,18,19^. Alongside a depletion of lung resident alveolar macrophages, there is a concurrent accumulation of pro-inflammatory mononuclear phagocytes which are peripherally recruited given correlation with cells in corresponding peripheral blood samples^34^. Macrophage populations expressing pro-fibrotic genes similar to those found in idiopathic pulmonary fibrosis also accumulate^35^. We observed progressive infiltration of both an inflammatory subset of monocyte/macrophages (IL1R^HI^, IL6R^HI^ and HLA-DR^HI^) as well as a CD206^HI^ monocyte/macrophage subset most likely to represent tissue repair macrophages with M2 polarisation^36^ as DAD progresses. The inflammatory group increased their interactions with interferon-responsive neutrophils and the repair group with T lymphocytes early in disease progression, prior to overt tissue damage. These early interactions may be critical in establishing dysregulated inflammatory responses and the over-zealous, deleterious repair processes which seem to be driven by macrophages. Proactive targeted modification of such interactions at disease detection may have some clinical application, if not for COVID-19 then for other viral illnesses which can lead to ARDS.

Similarly, our data shows that a second immune signature of COVID-19 DAD progression is a progressive infiltration of lymphocytes^18^. This rise was accounted for by naïve and memory CD4+ T cells, memory CD8+ T cells, B cells and plasma cells. T cells, similar to monocytes/macrophages are thought to have dual roles in COVID-19, from a protective response in mild to moderate disease to a dysregulated one in severe cases^37^. Lung resident and infiltrating B cells and plasma cells have received significantly less attention than T cells, although SARS-CoV-2-specific B cells have certainly been found in lung and lung-associated lymph nodal tissue^38^. Increased B lymphocytes in COVID-19 BALF correlate with evidence of severe disease^39^. Early recruitment and diapedesis of B lymphocytes/plasma cells is suggested from our results given their increased interaction with vascular endothelial cells in COVID positive compared to COVID negative tissue with preserved lung architecture. As DAD progresses however, there is continued plasma cell infiltration and increased B cell/plasma cell interactions with neutrophils and mononuclear phagocytes which may indicate a role in disease progression.

Lung tissue is inherently malleable and subject to both anatomic variation in inflation as well as variance in tissue preparation for histological sectioning. Whilst attempts at standardisation by post-mortem lung inflation have previously been described^40^, their routine use is impractical and unlikely to be accurate. Variability in alveolar filling is notable across disease processes such as COVID-19 where alveolar type II cell destruction results in reduced surfactant production and reduced surface tension; airspace occlusion by oedema/haemorrhage/fibrin balls/neutrophil extracellular traps; and increased connective tissue production and contraction^41^ can all contribute. Underpinning our data is the ability to quantify absolute and relative cellularity in the PMLT. However, the conventional measure of cellularity, cells/mm^2^ of section area, used is confounded by variations in the area of sections occupied by airspace both in diseased and healthy lung. We showed that COVID-19 DAD progression is characterised by significant airspace obliteration, as reported in prior literature^41^. Using the conventional metric of cells/mm^2^, alveolar epithelium and vascular endothelium increased throughout DAD. This would be unexpected given SARS-CoV-2 infect alveolar cells^28^, induce apoptosis^42^ and alveolar cell injury^33^ as well as observations of endothelialitis and endothelial apoptosis in COVID-19 PMLT^43^. However, application of an airspace correction factor showed a *decline* in alveolar epithelium and vascular endothelium as DAD progresses. This confirmed that using cells/mm^2^ is subject to an artefact from progressively obliterated airspaces and increased cellular tissue in the region of interest. To our knowledge, correcting for airspace is rarely used despite providing valuable information and should be considered for analysis of pliable tissue. We suspect it is most appropriately used in disease processes where airspace variability is secondary to primary airspace obliteration such as with progressive DAD rather than bronchopneumonia in which neutrophils invade the alveolar space. Another limitation of airspace correction lies is that non-cellular space is not exclusively air and includes other non-cellular material such as oedema.

This study had several additional limitations. Clinical data available varied due to collection across the three contributing institutions; a subset died in the community with limited information prior to death; time to post-mortem varied and a level of tissue degradation might have occurred; though our control tissue appeared histologically normal, it was obtained from deceased and therefore by definition unhealthy donors.

In summary, we have presented a comprehensive assessment of the cellular signature of COVID-19 DAD progression using a unique airspace correction method to normalise cell counts and account for a progressive march towards airspace obliteration. Sustained increases in MnPs and lymphocytes and a loss of AECs and endothelial cells are the hallmark of COVID-19 DAD progression and although neutrophils were overall stable, there is a shift in several functional subsets. Finally, we performed a neighbourhood analysis focussed on the distinct temporal phases of DAD progression and identified that critical immune cell interactions occur early in the disease process, prior to overt tissue damage.

## 6. Supplemental Figure Legends

**Figure S1: OPTIMAL segmentation achieves excellent segmentation of single cells across all 8 pathology types in COVID19 PM lung tissue:** Example images for each of the 8 pathology classes (as indicated) showing the results of OPTIMAL single cell segmentation as a probability mask from Ilastik (left panel image) and the subsequent cell boundary mask derived from CellProfiler segmentation (right image panel).

**Figure S2: Z-score normalisation of the arcsinh c.f. 1 transformed antibody-metal parameters removes significant batch effects in the data from staining and acquisition as well as biobank source.** A) PacMap plots of only arcsinh c.f.1 transformed parameter data (left plot, denoted “pre-correction”) and after a subsequent Z-score normalisation of the input parameters (right plot, denoted “post-correction”. Each plot is coloured by batch as shown in the associated figure legend. B) Batch-specific density-based PacMap plots derived from the fully corrected (arcsinh c.f.1 transformed and Z score normalised) parameter set. C) PacMap plots as in A but coloured by biobank source (as per legend). D) As per B but showing density-based PacMap plots per biobank source.

**Figure S3: There is no detectable SARS-CoV2 spike or capsid protein in any of the 7 pathology classes from infected samples as judged by positive and negative TMA controls.** A) Pseudo-coloured multichannel images from uninfected (left image panel) and infected (right image panel) FFPE-embedded vero cell pellet TMAs that were processed, stained and measured alongside cohort tissue samples. The overlaid signals are as per the legend and include the nuclear counterstain iridium (red) plus SARS-CoV2 spike (blue) and nucleocapsid (green). B) Single cell level (semi) quantitative data was derived using our OPTIMAL segmentation, feature extraction and exploration approach allowing us to determine the mean signal intensity of spike (upper panels) and nucleocapsid (lower panels) for each of the 8 pathology classes relative to our TMA positive and negative controls.

**Figure S4: A schematic of the Tier 1/Tier 2 analysis workflow.** A schematic breakdown of the analysis process used to identify and quantify the cell types and states at Tier1 and Tier 2 clustering levels.

**Figure S5: Differential analysis of the tier 1 cluster frequencies across all ROIs as a function of pathology class reveals key cell signatures.** A) Graphs showing the frequencies of key tier 1 immune cell types for each of the 8 pathology classes. From left to right; neutrophils, mononuclear phagocytes, T cells, B/Plasma Cells and NK Cells. B) Analogous graphs as shown in A but for major tier 1 structural cell types. From left to right; Platelets/Megakaryocytes, AT1, AT2, vascular endothelium and lymphatic endothelium. Differences between pathology classes were considered statistically significant at p<0.05.

**Figure S6: Differential and comparative analysis of the tier 1 cluster counts per mm^2^ of ROI versus per mm^2^ of lung tissue (adjusted) across all ROIs as a function of pathology class reveals the importance of factoring air gap loss in to cell counts.** A) Graphs showing the cell number per mm^2^ of ROI (upper panels) and for cell numbers adjusted to tissue area in mm^2^ (lower panels) for tier 1 immune cell types for each of the 8 pathology classes. From left to right; neutrophils, mononuclear phagocytes, T cells, B/Plasma Cells and NK Cells. B) As in A but showing data for structural cell types. From left to right; Platelets/Megakaryocytes, AT1, AT2, vascular endothelium and lymphatic endothelium. Differences between pathology classes were considered statistically significant at p<0.05.

**Figure S7: Subgroup analyses performed comparing sex, ethnicity and pandemic wave.** (A) PacMap dimensionality reduction plots coloured by patient sex, ethnicity and pandemic wave as shown in the corresponding legends. (B) Differential analysis plots of the same clinical metadata as in A showing the numbers of indicated cell types as a function of total tissue (airspace loss-corrected counts per mm^2^).

**Figure S8: Differential analysis of the tier 2 cluster frequencies across all ROIs as a function of pathology class reveals key cell signatures.** Graphs showing the frequencies of all 25 tier 2 clusters for each of the 8 pathology classes. Differences between pathology classes were considered statistically significant at p<0.05.

**Figure S9: Differential analysis of the tier 2 cluster counts per mm^2^ of ROI as a function of pathology class reveals key cell signatures.** Graphs showing the cell counts per mm^2^ of ROI for all 25 tier 2 clusters for each of the 8 pathology classes. Differences between pathology classes were considered statistically significant at p<0.05.

**Figure S10: Differential analysis of the tier 2 cluster counts per mm^2^ of lung tissue as a function of pathology class reveals key cell signatures.** Graphs showing the cell counts per mm^2^ of lung tissue for all 25 tier 2 clusters for each of the 8 pathology classes. Differences between pathology classes were considered statistically significant at p<0.05.

**Figure S11: Spatial neighbourhood analysis of tier 1 clusters for all 8 pathology classes.** Heat maps for all of the 8 pathology classes involved in classical and divergent disease progression showing the significance of interaction, avoidance or indifference for all 10 tier 1 clusters as per the legend. Red dotted squares are shown to aid with identifying and interpreting key interactions or avoidances as per the legend key.

**Figure S12: Spatial neighbourhood analysis of tier 2 clusters for all 8 pathology classes.** Heat maps for all of the 8 pathology classes involved in classical and divergent disease progression showing the significance of interaction, avoidance or indifference for all 25 tier 2 clusters as per the legend. Red dotted squares are shown to aid with identifying and interpreting key interactions or avoidances as per the legend key.

## 7. Supplemental Methods

**Supp Methods 1: Tissue sources and corresponding ethics approval**

Human samples used in this research project were partly obtained from the Newcastle Hospitals CEPA Biobank and their use in research is covered by Newcastle Hospitals CEPA Biobank ethics – REC 17/NE/0070. Additional human samples used in this research project were obtained from the Imperial College Healthcare Tissue Bank (ICHTB). ICHTB is supported by the National Institute for Health Research (NIHR) Biomedical Research Centre based at Imperial College Healthcare NHS Trust and Imperial College London. ICHTB is approved by Wales REC3 to release human material for research (22/WA/0214). Additional human samples used in this research project were obtained from the ICECAP tissue bank of the University of Edinburgh. ICECAP is approved by the East of Scotland Research Ethics Service to release human material for research (16/ED/0084).

**Supp Methods 2: Manufacture of Control Tissue MicroArray (TMA) Material**

To provide positive and negative staining controls for all 40 antibodies in our panel as well as provide empirical controls for batch effects we prepared FFPE Tissue MicroArrays (TMAs) blocks, composed of human tonsil tissue, as well as both SARS-CoV2-infected (BetaCoV/England/2/2020, obtained from the UK Health Security Agency)) and uninfected Vero E6 cells. SARS-CoV-2 is a Hazard Group 3 pathogen (Advisory Committee on Dangerous Pathogens, UK), as such infections were performed in a dedicated Containment Level 3 (CL3) facility by trained personnel as described in Hatton *et al.* (2021)^44^. Vero E6 cells were seeded in a 175ml flask until 90% confluent then infected with 1.5×10^6 PFU/mL of SARS-CoV-2 diluted in 2% FCS DMEM. The inoculum was removed after 2 hours and replaced with 30mL of warm 2% FCS DMEM. After 72 hours, supernatant was collected and centrifuged at 2000rpm for 30 minutes at 4’C. Supernatant was removed and the pellet was then resuspended in 4% formaldehyde for 1 hour at RT. Cells were then spun at 500g for 5 mins and resuspended in 70% alcohol solution/IMS. Cell pellets were processed and paraffin embedded at the Novopath Research Service (NovoPath, Department of Pathology, Newcastle Hospitals NHS Foundation Trust, Newcastle upon Tyne, UK). 2mm cores from paraffin embedded SARS-CoV-2 infected Vero E6 cells were embedded alongside 2mm cores of uninfected Vero E6 cells and Tonsil tissue, to produce a control tissue microarray block that was mounted on super frost slides and processed/stained alongside each batch of patient samples.

**Supp Methods 3: Conjugation and validation of metal-tagged antibodies**

Metal tags were paired with antibodies based on the relative staining intensity of each marker as determined by IF following the rules of “best practice” for CyTOF panel design^45^ using the Maxpar X8 metal conjugation kit following manufacturer’s protocol (Standard Biotools, CAT#201300). Antibodies conjugated to platinum isotopes 194 Pt and 198 Pt were conjugated as described in Mei *et al*. 2015 ^46^. Conjugation success was determined by measuring antibody recovery post-conjugation, metal addition by binding the antibody to iridium labelled antibody capture beads AbC™ Total Antibody Compensation Beads (Thermo Fisher, USA, CAT#A10513) and acquiring on a Helios system (Standard Bio-tools, USA), and finally a retained ability to recognise antigen post-conjugation using either a two layer IF or directly by IMC using the Hyperion imaging module (Standard Bio-Tool) connected to the Helios.

**Supp Methods 4: IMC set up, validation and ROI ablation**

Prior to each slide acquisition, the Hyperion Tissue Imager was calibrated/QC’d to achieve reproducible sensitivity based on the detection of 175Lutetium by ablating a single multi-element-coated “tuning slide” (Standard Biotools, USA) using the manufacturer’s “auto tune” application. After tuning, TMA control and experimental slides were loaded onto the Hyperion system to create Epi-fluorescence panorama images of the tissue surface and regions of interest (ROI) were set based on OMERO annotations. A small region of tonsil tissue was targeted to test that the chosen laser power was able to ablate the entire tissue depth. First, three 0.25mm^2^ ROIs, one per TMA control, were ablated followed by ROIs from the post-mortem cases with ROI sizes ranging from 0.25 – 1 mm^2^. Ablations were performed at 200Hz laser frequency to create a resultant MCD file containing all data from all ROIs for a given slide/case. MCD files were then opened in MCD Viewer software (Standard Bio-Tools) to perform a qualitative, visual QC of the staining intensity and pattern with the initial IF images as a benchmark. All images were exported as 16-bit single multi-level TIFFs using the “export” function from the “file” menu. For efficiency, all open collection channels from the experimental acquisition template (in this case, 60, including several “Blank” channels for QC purposes) from all ROIs were left selected and any image/channel removal was dealt with later in the analysis. The multi-level 16-bit TIFF images were then input into our OPTIMAL pipeline^22^ for data exploration at the single cell, spatial level.

## Data Availability

All data produced in the present study are available upon reasonable request to the authors

## Author Contributions

**Luke Milross MD**: conceptualization, formal analysis, investigation, data curation, writing – original draft, visualisation; **Bethany Hunter**: methodology, validation, formal analysis, investigation, data curation, writing – original draft, visualisation; **David McDonald PhD**: formal analysis, investigation, data curation, writing – review & editing; **George Merces PhD**: methodology, software, formal analysis, data curation, writing – review & editing; **Amanda Thompson**: validation, writing – review & editing; **Catharien M.U. Hilkens**: data curation, writing – review & editing; **John Wills PhD**: software, formal analysis, writing – review & editing; **Paul Rees PhD**: software, formal analysis, writing – review & editing; **Kasim Jiwa**: resources, writing – review & editing; **Nigel Cooper**: resources, writing – review & editing; **Joaquim Majo**: cenceptualization, methodology, formal analysis, data curation, writing – review & editing; **Helen Ashwin**: data curation, writing – review & editing; **Christopher JA Duncan DPhil**: resources, validation, writing – review & editing; **Paul M Kaye PhD**: conceptualization, methodology, writing – review & editing; **Omer Bayraktar PhD**: conceptualization, methodology, writing – review & editing; **Andrew Filby PhD**: conceptualization, methodology, software, validation, formal analysis, investigation, resources, data curation, writing – original draft, writing – review & editing, visualisation, supervision, project administration; and **Andrew J. Fisher PhD FRCP**: conceptualization, methodology, formal analysis, resources, writing – original draft, writing – review & editing, supervision, project administration.

## Acknowledgements

The authors would like to acknowledge the tissue donors and their families for their contribution to medical science, NOVOPATH for the supply and preparation of the tissue used in this study and other members of the NU-FCCF and BioImaging Unit for helpful discussions. Finally we thank Catherine F. Hatton and Jarmila S. Spegarova for providing SARS-CoV-2 infected material as well as the NU-Infectious Diseases Facility (IDF) for providing an excellent and approved environment to work at level three pathogen containment.

