## Supplemental Tables and Figures for "Distinct lung cell signatures define the temporal evolution of diffuse alveolar damage in fatal COVID-19"

**Supp Table 1.** *Characteristics of the cohort*

| Cohort Characteristics – n=40 | |
|---|---|
| **Age** | |
| Range | 22-98 |
| Median | 72 |
| **Sex** | |
| Female | 8 (20%) |
| Male | 32 (80%) |
| **Ethnicity** | |
| BAME | 8 (20%) |
| White | 32 (80%) |
| **Place of death** | |
| Community | 12 (30%) |
| Hospital | 28 (70%) |
| **Pandemic Wave** | |
| First | 30 (75%) |
| Second | 10 (25%) |
| **Pre-existing comorbidities** | |
| Cardiac disease | 15 (37.5%) |
| Hypertension | 9 (22.5%) |
| Diabetes mellitus | 8 (20%) |
| COPD/Asthma | 13 (32.5%) |
| CKD | 6 (15%) |
| Smoking history | 9 (27.5%) |
| Obesity | 11 (27.5%) |
| Dementia | 7 (17.5%) |

**Supp Table 2.** *Disease characteristics of the cohort*

| Disease Course | |
|---|---|
| **Duration of Illness (n=40)** | |
| ≤15 days | 21 (52.5%) |
| >15 days | 14 (35%) |
| Unknown | 5 (12.5%) |
| **Symptoms (n= 40)** | |
| Respiratory | 30 (75%) |
| Enteric | 7 (17.5%) |
| Fever | 22 (55%) |
| Systemic | 23 (57.5%) |
| Unknown | 3 (7.5%) |
| **Exposure to medications** | |
| Corticosteroids (n=38) | 10 (26.3%) |
| Antibiotics (n=39) | 25 (64.1%) |
| Anticoagulation (n=37) | 20 (54.1%) |

**Supp Table 3.** *Regions of interest selected for each histological pattern*

| Predominant Histology Phenotypes (n=40) | |
|---|---|
| EDAD | 128 |
| MDAD | 25 |
| ODAD | 60 |
| BRON | 45 |
| PO-ACF | 19 (from 2 cases) |
| IPM | 8 (from 1 case) |
| PRESneg | 30 |
| PRESpos | 24 |

**Supp Table 4.** *Histological Criteria utilised for pathology phenotype identification and ROI selection*

| Pathology Code | Hallmark Characteristic | Supportive Features |
|---|---|---|
| EDAD | Presence of hyaline membranes (HM)<br>Absence of fibroblastic organisation (FO) | Mild interstitial inflammation<br>Type II pneumocyte hyperplasia<br>Alveolar haemorrhage |
| ODAD | Presence of FO<br>Absence of HM | Fibroblast aggregates<br>Scattered inflammatory cells<br>Collagen deposition<br>Squamous metaplasia |
| MDAD | Presence of HM and FO in any degree | |
| BRON | Neutrophilic exudate | +/- tissue damage<br>Alveolar haemorrhage |
| PO-ACF | Alveolar / interstitial oedema<br>Absence of tissue injury | Pulmonary vascular congestion |
| IPM | Identification of fungal hyphae<br>Tissue inflammation / necrosis | Angioinvasion<br>Positive culture |

**Supp Table 5.** *Antibody panel details*

| Target | Clone | Vendor | Catalogue # | Metal | Working Concentration (ug/mL) | Major target/cell type/population | Main spatial location |
|---|---|---|---|---|---|---|---|
| SARS-CoV-2 Spike | E7M5X | Cell Signalling Technology | 42172S | 113In | 5 | SARS-CoV-2 virus, Spike protein | Various |
| CD45RO | UCHL1 | Thermofisher | 14-0457-82 | 115In | 4 | Memory T cells (+ macrophages) | Immune cell marker, surface |
| CD45RA | HI100 | Thermofisher | 14-0458-82 | 141Pr | 4 | Naïve T cells | Immune cell marker, surface |
| CD68 | KP1 | Biolegend | 372902 | 142Nd | 5 | Tissue macrophages (+ monocytes and monocyte-derived dendritic cells) | Immune cell marker, surface |
| CD8a | C8/144B | Biolegend | 372902 | 143Nd | 6 | Cytotoxic T cells | Immune cell marker, surface |
| KI67 | Polyclonal | Novus | NB500-170 | 144Nd | 3 | Proliferating cells | Non-specific functional marker, nuclear |
| Collagen I | 3D5E8 | Protein Tech | 66761-1-Ig | 145Nd | 2.5 | Connective tissues including extracellular matrix | Structural, extracellular |
| CD138 | 4F3A8 | Protein Tech | 67155-1-Ig | 146Nd | 3.5 | Plasma cells or precursors to plasma cells (plasmablasts) | Immune cell marker, surface |
| CD163 | EDHu-1 | BioRad | MCA1853 | 147Sm | 10 | Monocytes/macrophages and monocyte-derived dendritic cells | Immune cell marker, surface |
| IL1-R | Polyclonal | Thermofisher | PA5-28834 | 148Nd | 3.5 | Multiple cell types | Cell surface |
| CD42b | SP219 | Abcam | ab240268 | 149Sm | 8 | Platelets, megakaryocytes | Platelet cell surface, cytosolic in megakaryocytes. |
| MPO | 4C11F6 | Protein Tech | 66177-1-Ig | 150Nd | 5 | Polymorphonuclear leucocytes/granulocytes (neutrophils, basophils, eosinophils), monocytic cells. | Immune cell marker, cytosolic (lysosome) |
| SAR-Cov-2 Capsid | E2M5O | Cell Signalling Technology | 33717S | 151Eu | 5 | SARS-CoV-2 virus, Capsid protein | Various |
| B7 | TBC | Gifted Paul Morgan | NA | 152Sm | 20 | Antigen presenting cells | Cell surface, functional marker |
| CD56 | E7X9M | Cell Signalling Technology | 99746BF | 153Eu | 12 | Natural killer cells (+*alpha beta T cells, gamma delta T cells, dendritic cells, and monocytes*) | Immune cell marker, surface |
| Podoplanin | NZ-1.3 | Thermofisher | 14-9381-82 | 154Sm | 6 | Lymphatic endothelium | Structural, cell surface |
| CD69 | 15B5G2 | Novus | NBP2-25236 | 155Gd | 0.25 | Tissue-restricted T cells, NK cells, other immune cells, and platelets. | Immune cell marker, surface |
| EPCAM | Polyclonal | Abcam | ab71916 | 156Gd | 1 | Most epithelial cells (such as airway epithelial cells and pneumocytes) | Cell surface, particularly basolateral surface of epithelial cells |
| CD206 | 2A6A10 | Abcam | 60143-1-Ig | 157Gd | 0.5 | Macrophages and dendritic cells, associated with M2 macrophage polarisation | Immune cell marker, surface |
| CD79a | EP3618 | Abcam | ab239891 | 158Gd | 7.5 | B cells | Immune cell marker, surface |
| STING | D2P2F | Cell Signalling Technology | 13647 | 159Tb | 5 | Ubiquitous, IFN responding cells | Ubiquitous. Cell membrane and endoplasmic reticulum |
| TMPRSS | Polyclonal | Thermofisher | PA5-14265 | 160Gd | 10 | Expressed by multiple human cell types including but not limited to airway epithelial cells and pneumocytes. Facilitator of SARS-CoV-2 viral entry. | Cell surface |
| AQP5 | Polyclonal | Thermofisher | PA5-14264 | 161Dy | 5 | Type I pneumocytes (and other cell types including multiple immune cells) | Structural - Cell surface marker for Type I pneumocytes, apical membrane distribution |
| CD1c | 2A7C11 | Novus | NBP2-61726 | 162Dy | 2.5 | Dendritic cells, monocytes and monocyte-derived dendritic cells (and *B cells*) | Immune cell marker, surface |
| IFITM3 | Polyclonal | Protein Tech | 11714-1-AP | 163Dy | 5 | Ubiquitous, IFN responding cells | Non-specific functional marker, cell membrane and endolysosomal |
| ACE2 | 2F12A4 | Protein Tech | 66699-1-Ig | 164Dy | 5 | Expressed by multiple human cell types including but not limited to airway epithelial cells and pneumocytes. Facilitator of SARS-CoV-2 viral entry. | Cell surface |
| CD57 | HNK-1 | Biolegend | 359602 | 165Ho | 10 | T cells, NK cells | Cell surface, functional marker |
| p16 | EPR1473 | Abcam | ab186932 | 166Er | 10 | Ubiquitous, marker of cell senescence | Cytosolic/Nuclear |
| IL6-R | Polyclonal | Thermofisher | PA5-100836 | 167Er | 10 | Multiple cell types | Cell surface |
| Cleaved Caspase-3 | Asp175 | Cell Signalling Technology | 9579S | 168Er | 5 | Apoptotic cells | Non-specific functional marker, cytoplasmic |
| CD61 | D7X3P | Cell Signalling Technology | 13166BF | 169Tm | 5 | Megakaryocytes, platelets | Cell surface |
| CD3 | Polyclonal | Fluidigm | 3170019D | 170Er | 7.5 | T cells | Immune cell marker, surface |
| ProSPC | Polyclonal | Novus | NBP1-60117 | 171Yb | 4 | Type II pneumocytes | Cytoplasmic |
| CD31 | EPR3094 | Abcam | ab207090 | 172Yb | 2 | Platelets, endothelial cells (and *leukocytes*) | Cell surface |
| C3-30 | | Gifted Claire Harris | | 173Yb | 8 | Complement component C3 (activated complement) | Extracellular/cell membrane/ubiquitous |
| CD4 | EPR6855 | Abcam | ab181724 | 174Yb | 6 | T helper cells (*and NK T cells, innate lymphoid cells and macrophages*). | Immune cell marker, surface |
| HLA-DR | LN3 | Thermofisher | 14-9956-82 | 175Lu | 5 | 'Professional' APCs including B cells, DCs, macrophages, monocytes | Immune cell marker, surface |
| CD169 | SP213 | Abcam | ab245735 | 176Yb | 2.5 | Macrophages (and dendritic cells) | Immune cell marker, surface |
| CD147 | E1S1V | Cell Signalling Technologies | 13287BF | 194Pt | 4 | Widely expressed in multiple human cell types including leucocytes, epithelial cells and endothelial cells. Alternate facilitator of SARS-CoV-2 viral entry. | Cell surface |
| Beta-2-microglobulin | EPR21752-214 | Abcam | ab237032 | 198Pt | 1.5 | All nucleated cells | Cell surface |

Suppl. Figure 1

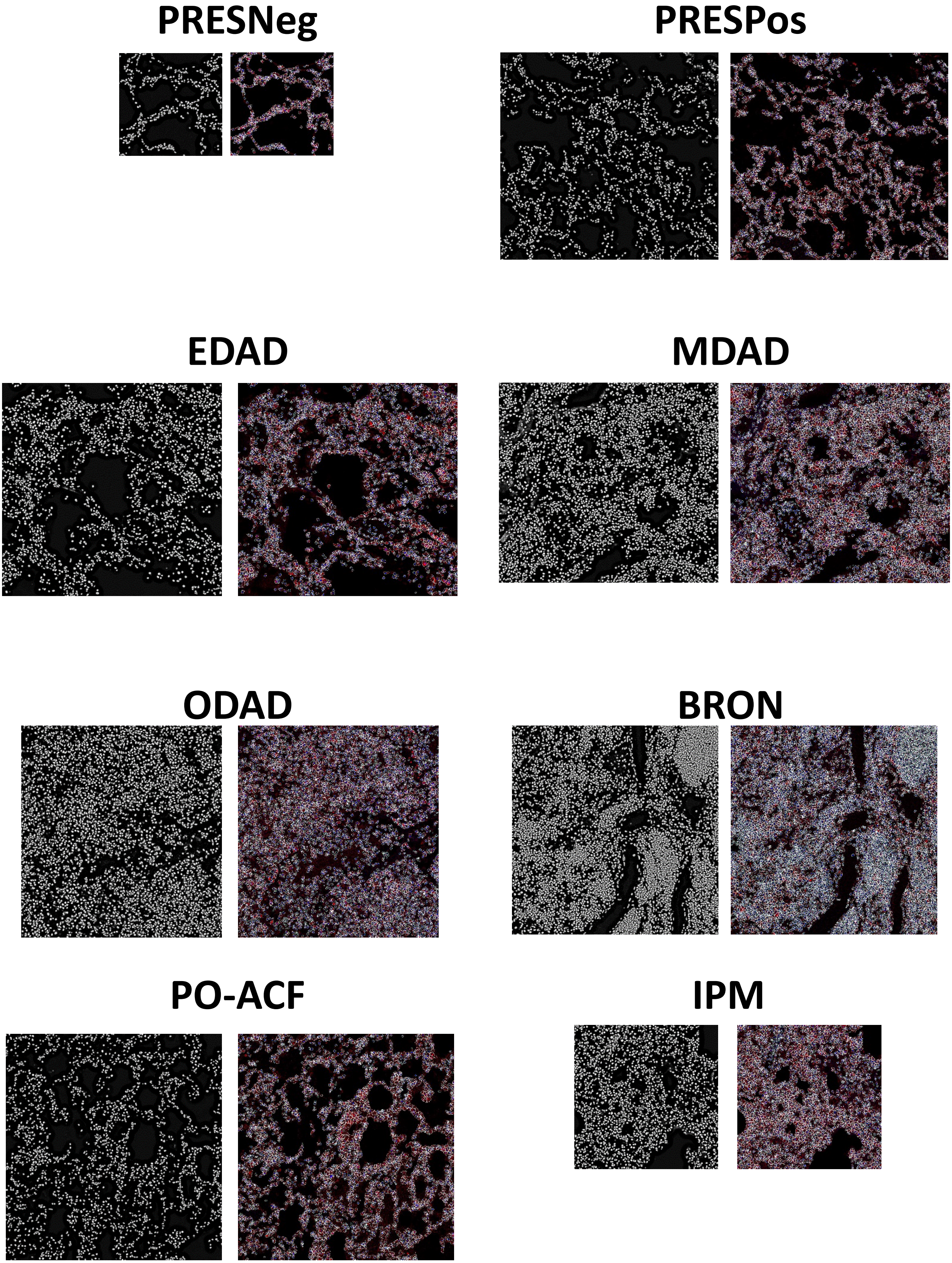

### Suppl. Figure 2

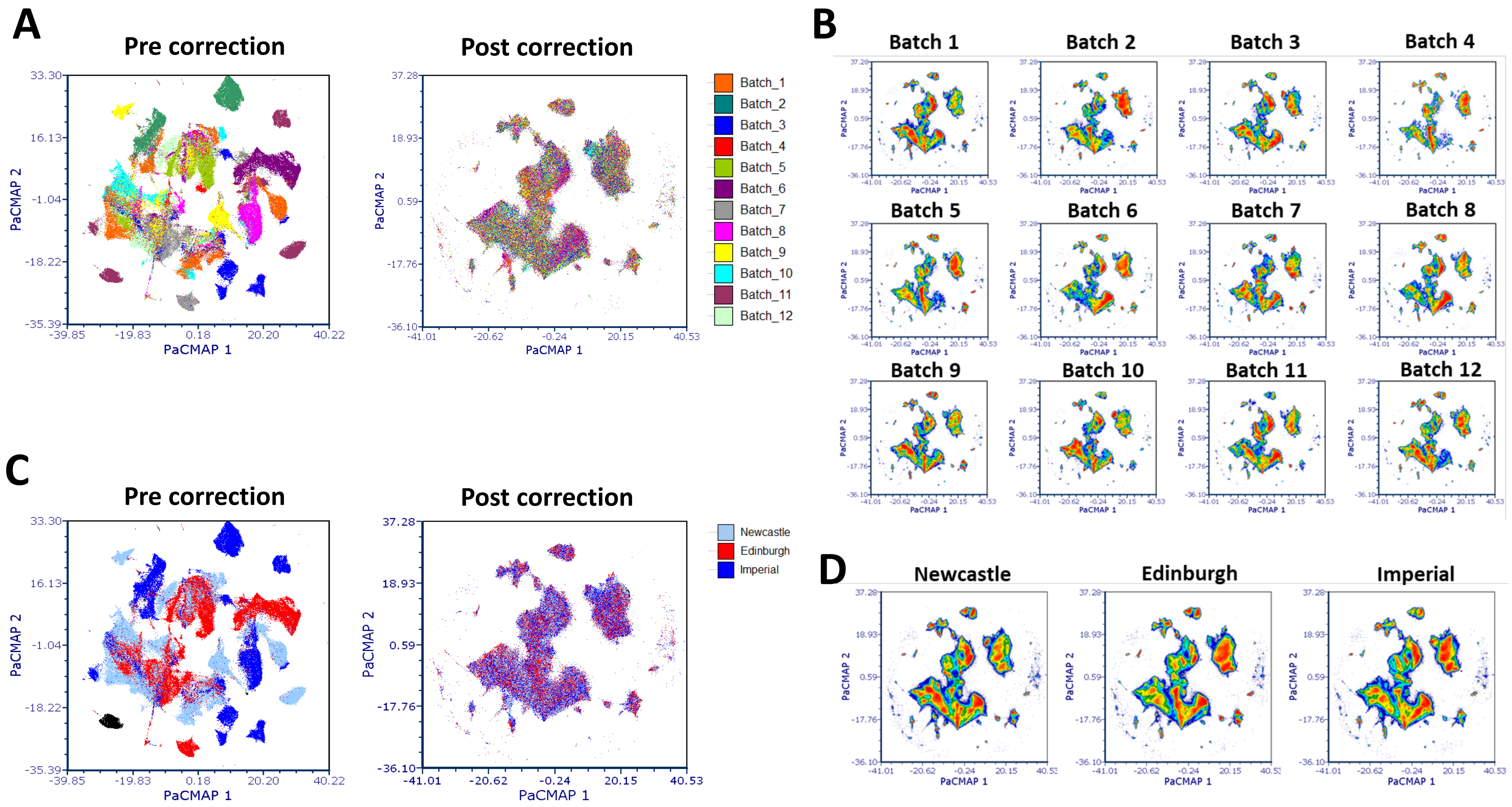

### Suppl. Figure 3

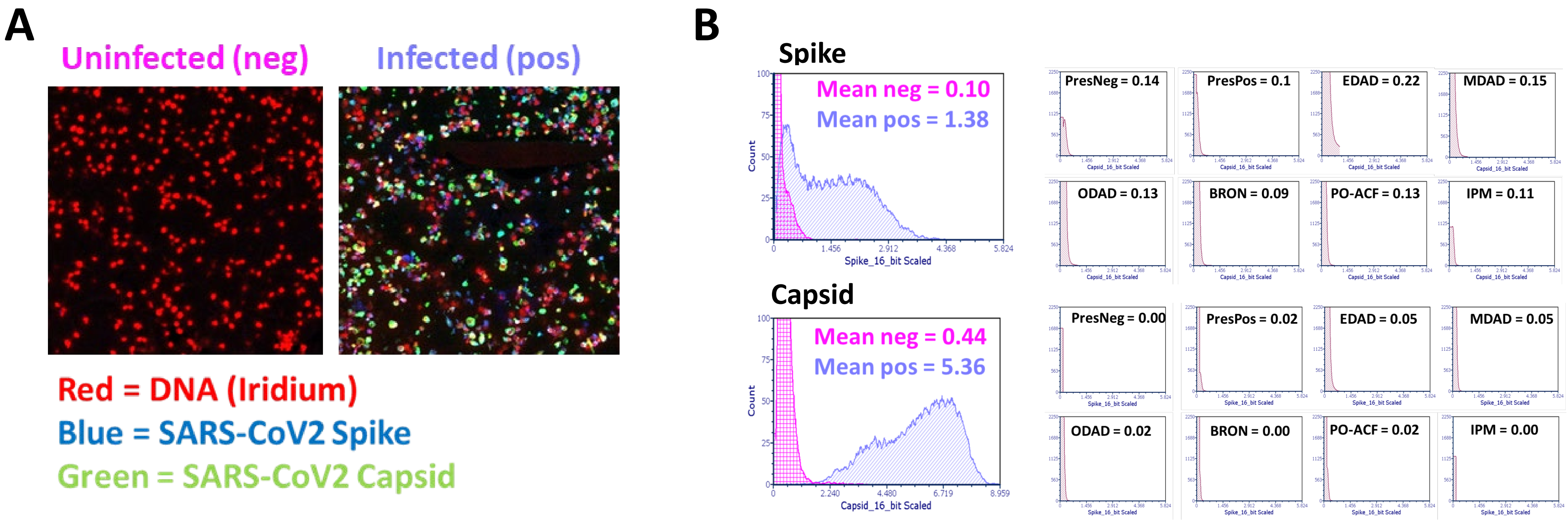

### Suppl. Figure 4

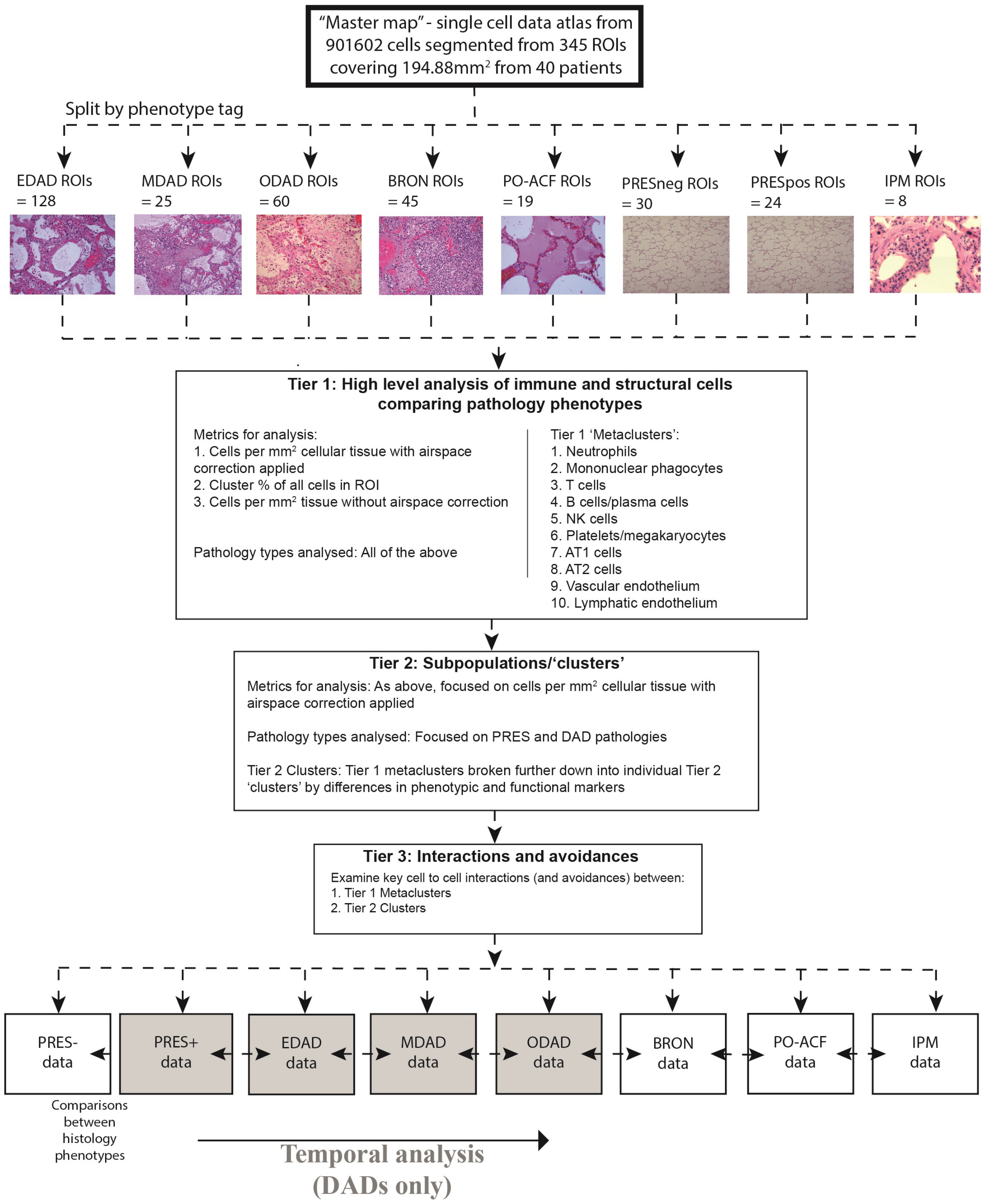

### Suppl. Figure 5

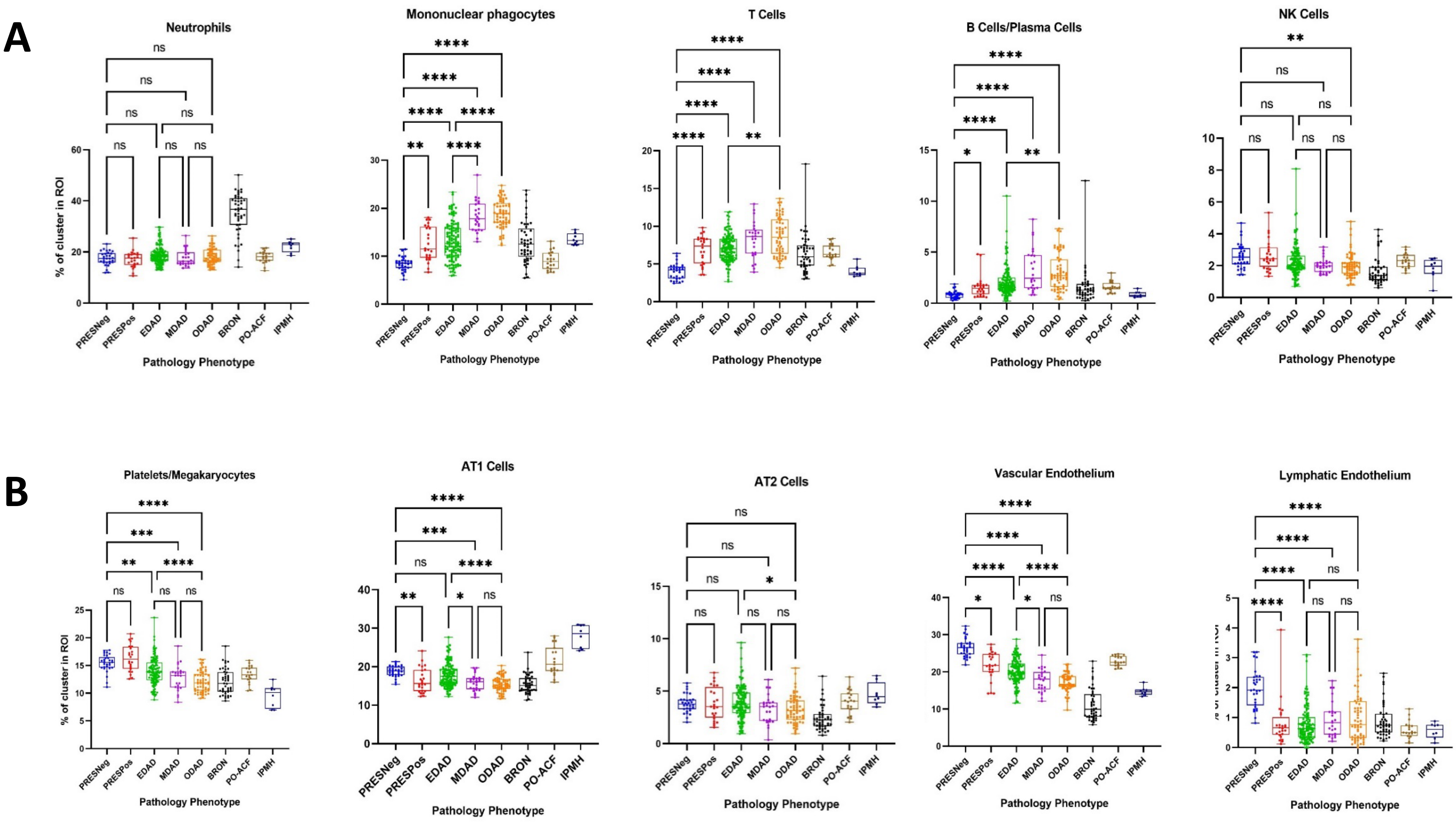

### Suppl. Figure 6

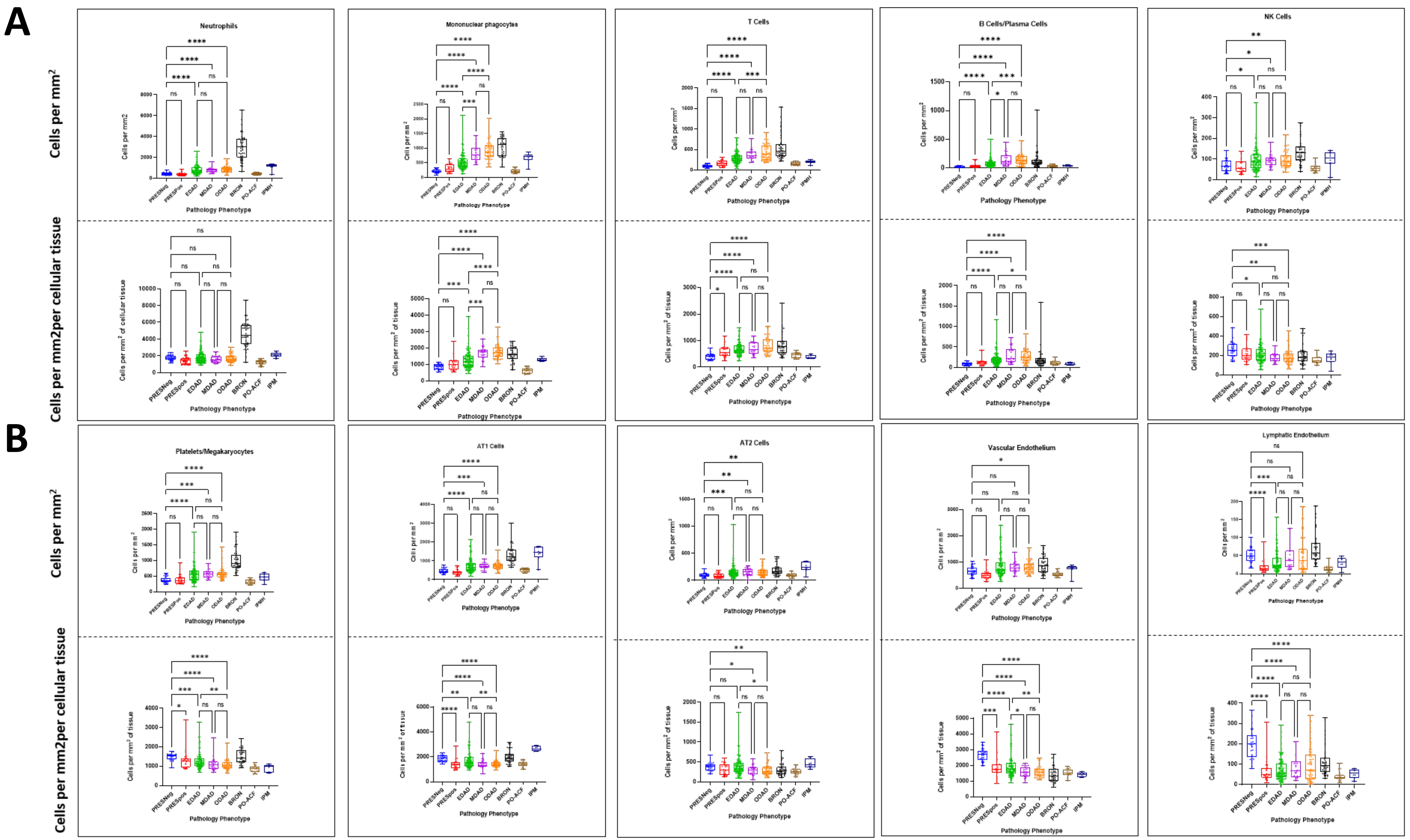

### Suppl. Figure 7

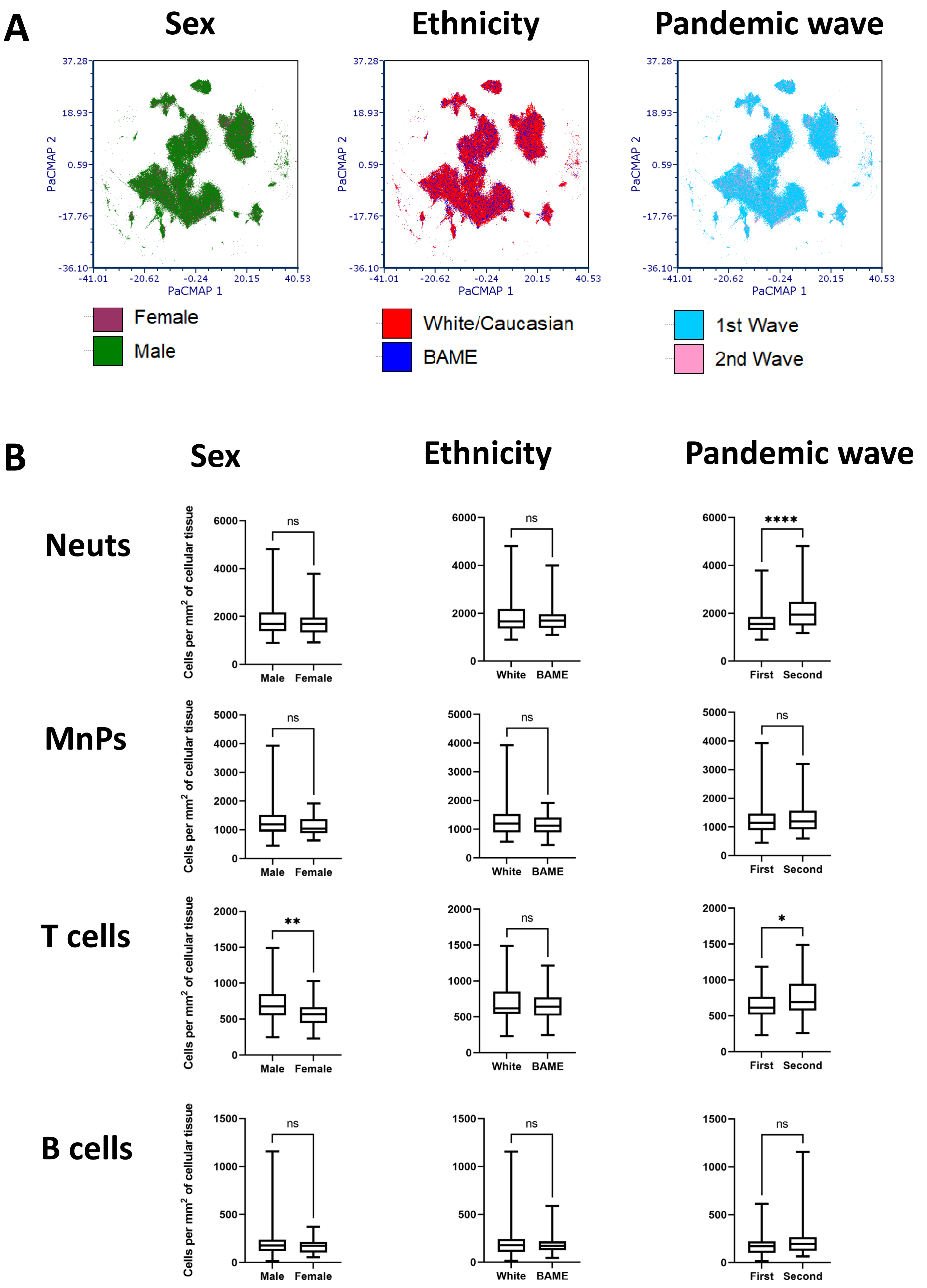

### Suppl. Figure 8

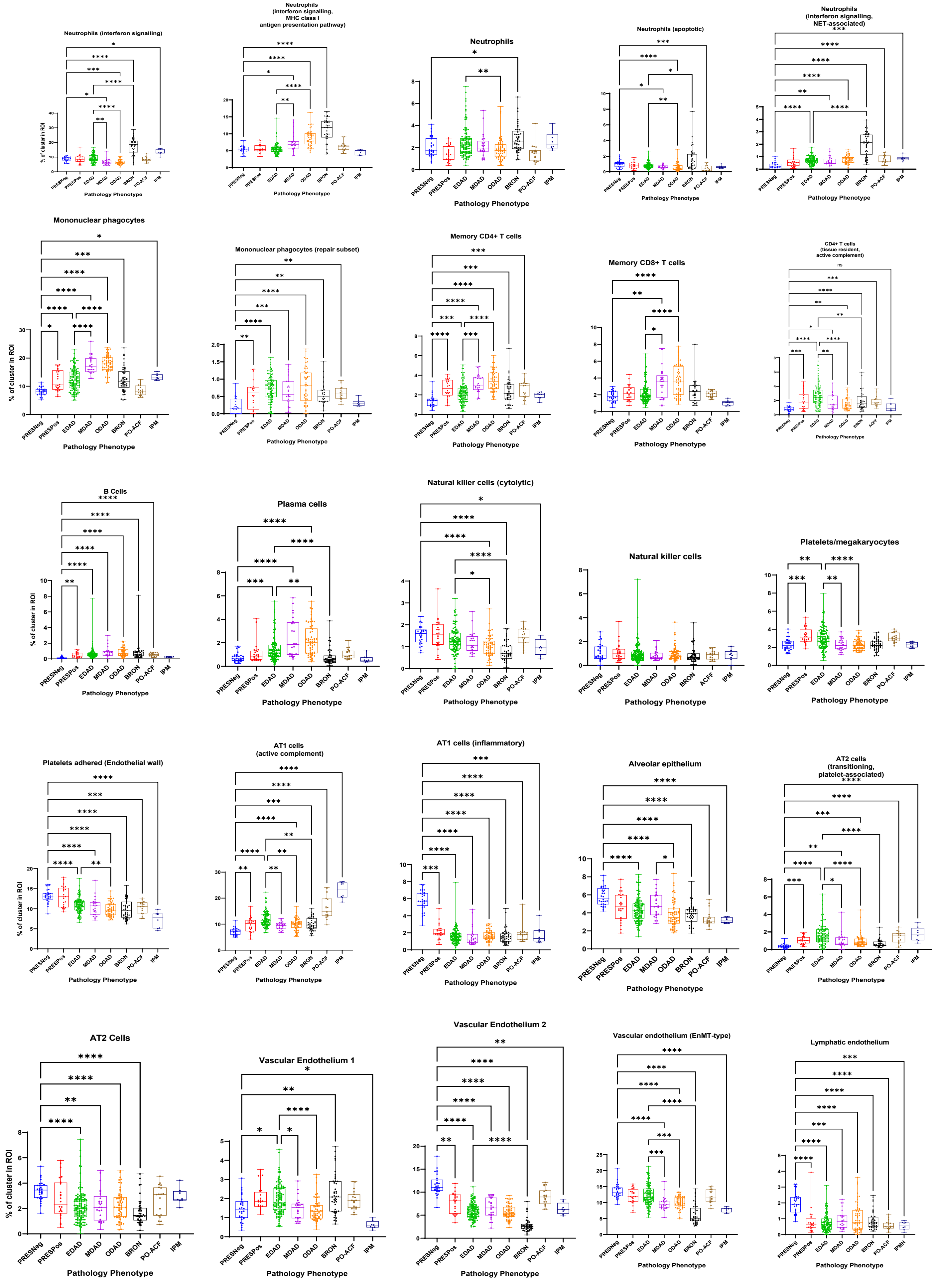

### Suppl. Figure 9

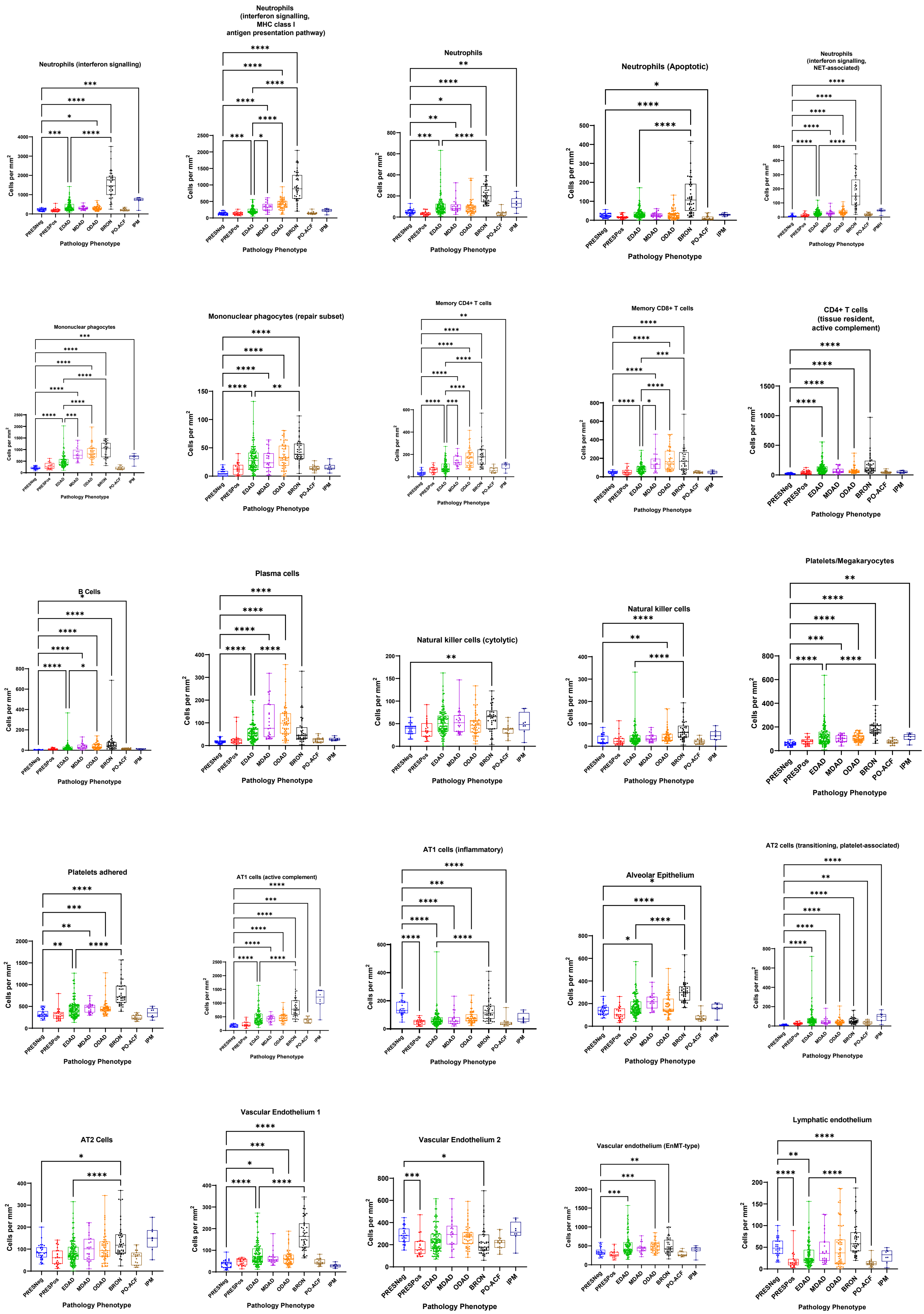

### Suppl. Figure 10

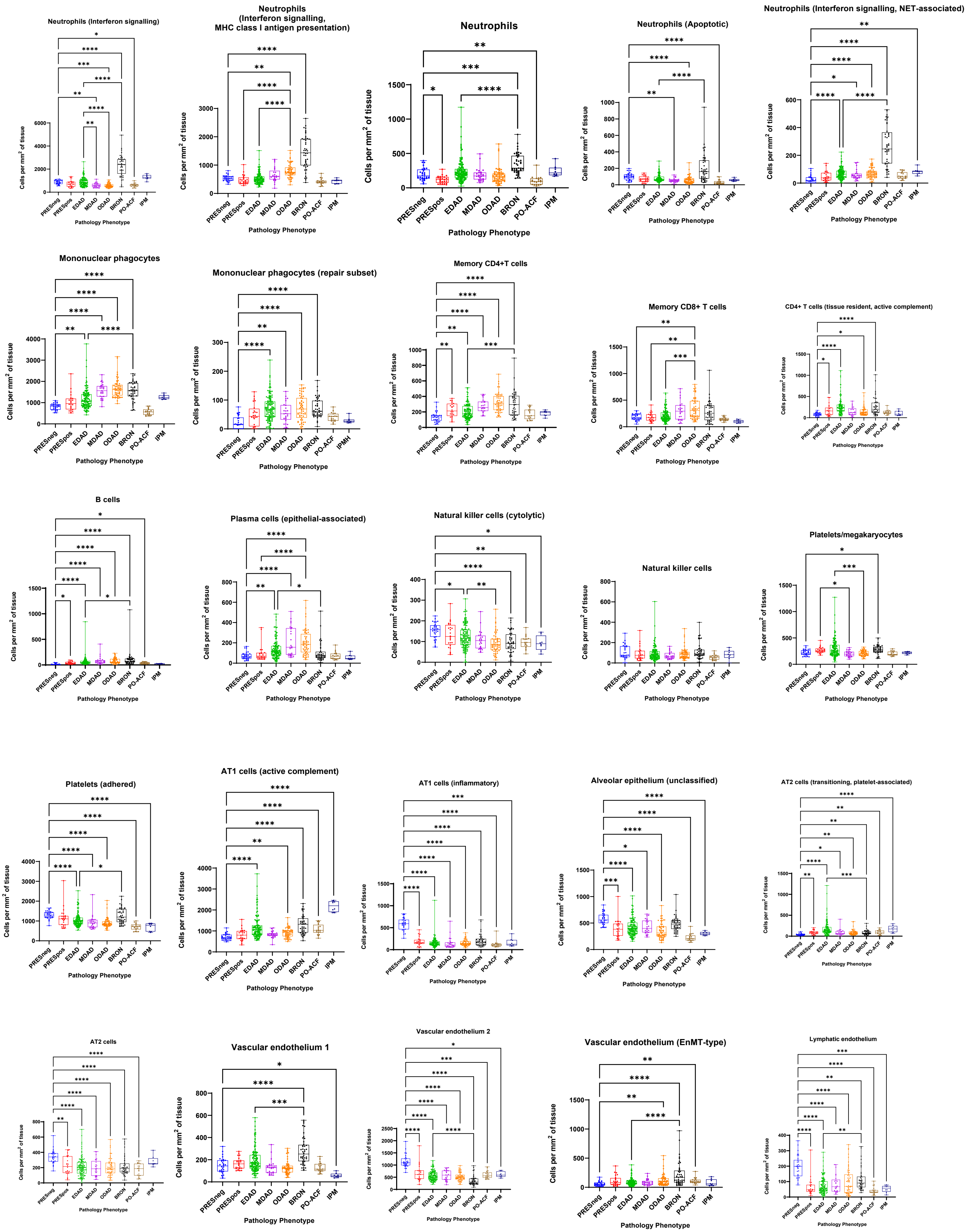

### Suppl. Figure 11

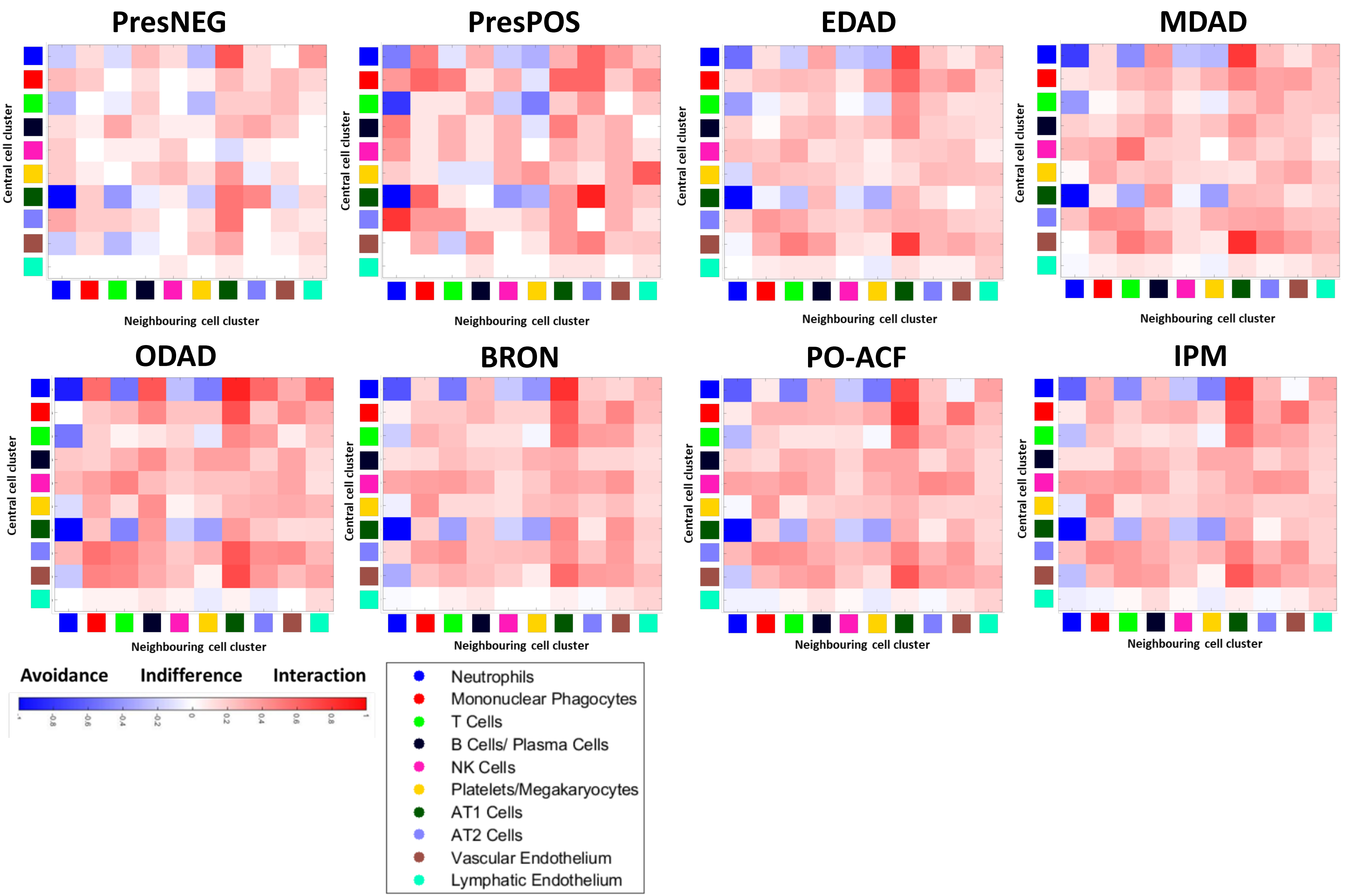

### Suppl. Figure 12

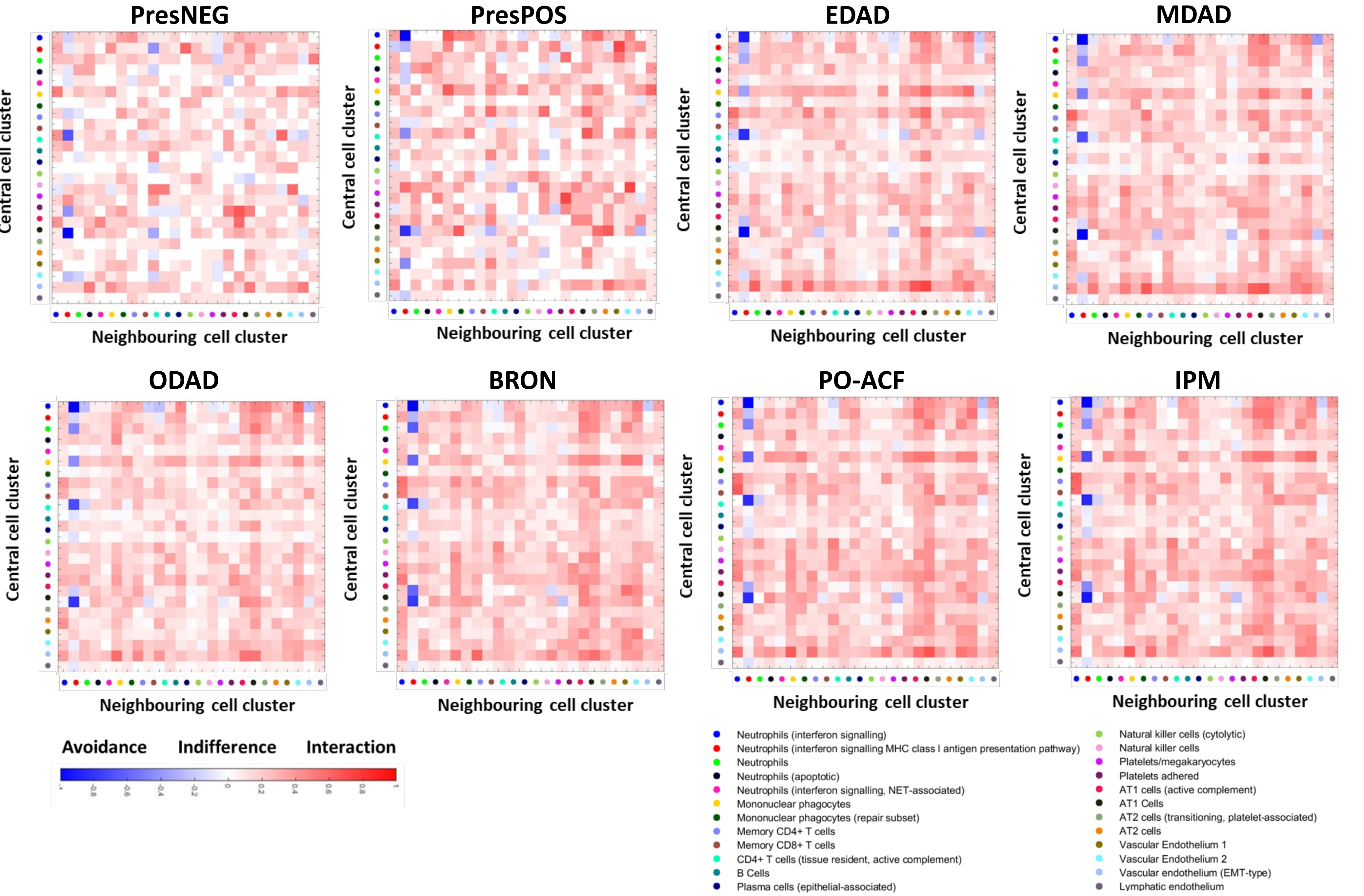
